# Cell-mediated and humoral immune response to SARS-CoV-2 vaccination and booster dose in immunosuppressed patients

**DOI:** 10.1101/2022.01.04.22268750

**Authors:** Lu M. Yang, Cristina Costales, Muthukumar Ramanathan, Philip L. Bulterys, Kanagavel Murugesan, Joseph Schroers-Martin, Ash A. Alizadeh, Scott D. Boyd, Janice M. Brown, Kari C. Nadeau, Sruti S. Nadimpalli, Aileen X. Wang, Stephan Busque, Benjamin A. Pinsky, Niaz Banaei

**Affiliations:** Department of Pathology, Stanford University School of Medicine, Stanford, CA 94305 USA; Department of Medicine, Division of Oncology, Stanford University School of Medicine, Stanford, CA 94305 USA; Sean N. Parker Center for Allergy & Asthma Research, Stanford, CA 94305 USA; Department of Medicine, Division of Infectious Diseases and Geographic Medicine, Stanford University School of Medicine, Stanford, CA 94305 USA; Department of Medicine, Division of Pulmonary, Allergy & Critical Care Medicine, Stanford University School of Medicine, Stanford, CA 94305 USA; Department of Pediatrics, Division of Pediatric Infectious Diseases, Stanford University School of Medicine, Stanford CA 94305 USA; Department of Medicine, Division of Nephrology, Stanford University School of Medicine, Stanford, CA 94305 USA; Department of Surgery, Division of Abdominal Transplantation, Stanford University School of Medicine, Stanford, CA 94305 USA; Clinical Microbiology Laboratory, Stanford Health Care, Palo Alto, CA 94304 USA

## Abstract

**Importance:** Data on the humoral and cellular immune response to primary and booster SARS-CoV-2 vaccination in immunosuppressed patients is limited.

**Objective:** To determine humoral and cellular response to primary and booster vaccination in immunosuppressed patients and identify variables associated with poor response.

**Design:** Retrospective observational cohort study.

**Setting:** Large healthcare system in Northern California.

**Participants:** This study included patients fully vaccinated against SARS-CoV-2 (mRNA-1273, BNT162b2, or Ad26.COV2.S) who underwent clinical testing for anti-SARS-SoV-2 S1 IgG ELISA (anti-S1 IgG) and SARS-CoV-2 interferon gamma release assay (IGRA) from January 1, 2021 through November 15, 2021. A cohort of 18 immunocompetent volunteer healthcare workers were included as reference. No participants had a prior diagnosis of SARS-CoV-2 infection.

**Exposure(s):** Immunosuppressive diseases and therapies.

**Main Outcome(s) and Measure(s):** Humoral and cellular SARS-CoV-2 vaccine response as measured by anti-S1 IgG and SARS-CoV-2 IGRA, respectively, after primary and booster vaccination.

**Results:** 496 patients (54% female; median age 50 years) were included in this study. Among immunosuppressed patients after primary vaccination, 62% (261/419) had positive anti-S1 IgG and 71% (277/389) had positive IGRA. After booster, 69% (81/118) had positive anti-S1 IgG and 73% (91/124) had positive IGRA. Immunosuppressive factors associated with low rates of humoral response after primary vaccination included anti-CD20 monoclonal antibodies (*P*<.001), sphingosine 1-phsophate (S1P) receptor modulators (*P*<.001), mycophenolate (*P*=.002), and B cell lymphoma (*P*=.004); those associated with low rates of cellular response included S1P receptor modulators (*P*<.001) and mycophenolate (*P*<.001). Of patients who responded poorly to primary vaccination, 16% (4/25) with hematologic malignancy or primary immunodeficiency developed a significantly increased humoral response after the booster dose, while 52% (14/27) with solid malignancy, solid organ transplantation, or autoimmune disease developed an increased response (*P*=.009). Only 5% (2/42) of immunosuppressed patients developed a significantly increased cellular response following the booster dose.

**Conclusions and Relevance:** Cellular vaccine response rates were higher than humoral response rates in immunosuppressed individuals after primary vaccination, particularly among those undergoing B cell targeting therapies. However, humoral response can be increased with booster vaccination, even in patients on B cell targeting therapies.

## INTRODUCTION

One of every 25 individuals in the U.S. is estimated to be immunocompromised^1^ and potentially at increased risk for severe COVID-19^2^ and breakthrough infections after vaccination^3^. Studies have demonstrated poor humoral response to SARS-CoV-2 vaccination in immunosuppressed patients^4,5^. On the other hand, cellular, or T cell vaccine responses appear to be less impaired in certain immunosuppressed groups, but are less well characterized^6–8^. Due to the decline in antibody titers over time after vaccination, booster shots have been recommended for all adults^9,10^. Boosters are thought to be especially important for immunosuppressed patients due to their impaired response to primary vaccination^11–13^. Here, we compare the humoral and cellular immune responses to SARS-CoV-2 after primary and booster vaccination among immunosuppressed patients. By retrospective analysis, we identify immunosuppressive factors that contribute to impaired response.

## METHODS

### Ethics Statement

This study was approved by the Stanford University Institutional Review Board (IRB-60171 and IRB-57519). Informed consent was obtained from volunteer healthcare workers before blood collection.

### Study Design

Assay design and interpretation for the anti-SARS-CoV-2 S1 IgG (anti-S1 IgG) assay, ACE2 blocking activity assay, and SARS-CoV-2 interferon gamma (IFN-γ) release assay (IGRA) are described in eMethods. We retrospectively assessed patients with IGRA ordered as part of clinical testing for patients at Stanford Health Care from January 1, 2021 through November 15, 2021. We also recorded available anti-S1 IgG antibody results. We performed two main analyses: primary vaccination response and booster response. Those patients with anti-S1 IgG or IGRA results collected at least 14 days following receipt of the second dose of either the Moderna mRNA-1273 or the Pfizer/BioNTech BNT162b2 mRNA vaccines or single dose of the Janssen Ad26.COV2.S vaccine were included in the primary vaccination analysis. One additional dose with any of the three vaccines was considered a booster dose. Patients with anti-S1 IgG or IGRA collected at least seven days following receipt of a booster dose were included in the booster analysis.

Electronic medical record (EMR) review was performed by one of five physician authors (NB, PB, CC, MR, LY) to collect data on underlying disease and active immune-suppressive/modulatory therapy (ISMT), including chemical drugs, biologics, and cellular therapy, such as hematopoietic stem cell transplant (HSCT) and CAR-T. Active ISMT was defined by the following conditions: receiving therapy 1) at the time of, or 2) up to four weeks prior (up to one year prior for HSCT and CAR-T) to administration of the first SARS-CoV-2 vaccine dose, or 3) anytime between administration of the first vaccine dose and the time of anti-S1 IgG and IGRA testing. ISMTs were additionally categorized by mechanism of action (eTables 1,2).

**Table 1.** Demographic information and assay results for cohorts after primary vaccination.

|  |  |  |  | Disease Categories |  |  |  |  |  |  |  |
| --- | --- | --- | --- | --- | --- | --- | --- | --- | --- | --- | --- |
|  | HCW<br>(n=18) | NISP<br>(n=28) | ISP<br>(n=427) | Active Heme<br>Malignancy<br>(n=29) | Inactive Heme<br>Malignancy<br>(n=65) | Primary<br>anemia<br>(n=9) | Solid Malignancy<br>(n=24) | Solid Organ<br>Transplant (n=67) | Autoimmune<br>Disease<br>(n=149) | 1°<br>Immuno-<br>deficiency<br>(n=42) | Multiple<br>Categories<br>(n=42) |
| On ISMT, % (n) | 0 (0) | 0 (0) | 69 (295) | 59 (17) | 45 (29) | 11 (1) | 46 (11) | 100 (67) | 89 (133) | 2 (1) | 86 (36) |
| Male:female ratio | 0.5:1 | 1.5:1 | 0.8:1 | 1.9:1 | 1.3:1 | 0.5:1 | 0.8:1 | 1.5:1 | 0.5:1 | 1.0:1 | 0.6:1 |
| Age, median (IQR), years | 43.5 (38.2-47.8) | 44.0 (35.8-64.2) | 49.0 (24.0-64.0) | 56.0 (29.0-70.0) | 39.0 (19.0-65.0) | 19.0 (16.0-24.0) | 20.5 (15.5-63.2) | 41.0 (17.0-59.5) | 53.0 (38.0-63.0) | 47.0 (32.8-65.5) | 57.5 (35.0-71.0) |
| IgG days since vaccine*, median (IQR) | 20.0 (17.0-21.0) | 158.0 (62.5-194.5) | 80.0 (41.0-127.0) | 48.0 (32.0-97.0) | 48.0 (32.8-95.8) | 78.0 (41.0-102.0) | 99.0 (53.2-129.5) | 66.0 (39.5-105.0) | 108.5 (62.8-152.2) | 81.0 (41.0-118.0) | 81.0 (45.5-122.5) |
| Anti-S1 IgG, median (IQR), OD ratio | 9.6 (9.3-9.9) | 5.7 (3.6-8.2) | 2.8 (0.2-7.1) | 0.7 (0.1-3.9) | 5.1 (0.3-8.4) | 8.5 (6.8-8.9) | 7.2 (4.9-8.7) | 3.3 (0.3-7.7) | 1.2 (0.2-5.2) | 3.5 (1.8-7.0) | 1.6 (0.2-6.1) |
| Anti-S1 IgG, % positive (n) | 100 (18/18) | 100 (27/27) | 62 (261/419) | 50 (14/28) | 69 (44/64) | 89 (8/9) | 88 (21/24) | 66 (44/67) | 53 (76/144) | 78 (32/41) | 52 (22/42) |
| ACE2 Blocking %, median (IQR) | 68.0 (50.0-90.8) | 12.0 (0.0-85.0) | 0.0 (0.0-43.8) | 0.0 (0.0-4.0) | 14.5 (0.0-94.8) | 100.0 (33.0-100.0) | 28.0 (0.0-85.5) | 0.5 (0.0-62.2) | 0.0 (0.0-10.2) | 0.0 (0.0-43.0) | 0.0 (0.0-22.0) |
| ACE2 Blocking, % positive (n) | 94 (17/18) | 44 (12/27) | 32 (134/419) | 18 (5/28) | 47 (30/64) | 78 (7/9) | 54 (13/24) | 34 (23/67) | 22 (31/144) | 34 (14/41) | 26 (11/42) |
| IGRA days since vaccine^, median (IQR) | 20.0 (17.0-21.0) | 185.0 (148.8-203.5) | 98.0 (56.0-142.0) | 86.5 (47.0-131.5) | 70.0 (41.8-121.2) | 78.0 (41.0-102.0) | 99.0 (60.2-137.2) | 81.0 (58.0-119.5) | 115.0 (78.0-156.0) | 83.5 (49.0-119.2) | 99.5 (64.0-147.5) |
| IFN-γ Response median (IQR), IU/mL | 1.3 (1.1-5.6) | 2.3 (1.4-4.3) | 1.9 (0.2-5.2) | 0.5 (0.1-4.4) | 2.8 (0.7-6.4) | 3.3 (3.0-8.3) | 4.6 (1.1-9.4) | 0.8 (0.1-2.3) | 2.2 (0.5-5.7) | 3.2 (0.4-7.0) | 0.4 (0.1-3.1) |
| IFN-γ Response, % positive (n) | 100 (18/18) | 92 (24/26) | 71 (277/389) | 58 (15/26) | 80 (45/56) | 100 (9/9) | 82 (18/22) | 58 (34/59) | 77 (106/137) | 75 (30/40) | 50 (20/40) |

Note: not all patients had both anti-S1 IgG and IGRA results available.
\*Number of days elapsed between primary vaccination and anti-S1 IgG testing. ^Number of days elapsed between primary vaccination and IGRA testing.

**Table 2.**
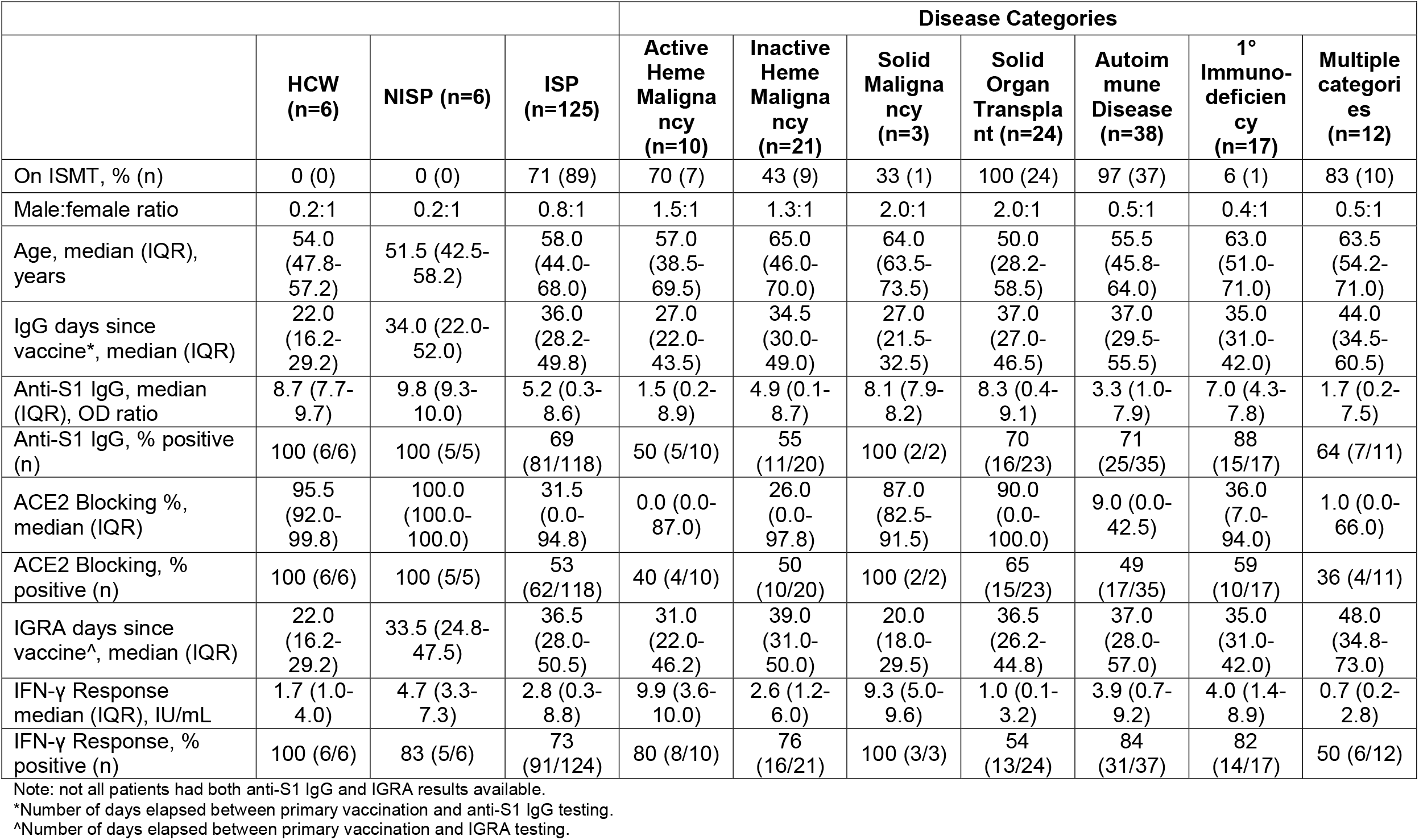
Demographic information and assay results for cohorts after booster vaccination.

|  |  |  |  | Disease Categories |  |  |  |  |  |  |
| --- | --- | --- | --- | --- | --- | --- | --- | --- | --- | --- |
|  | HCW<br>(n=6) | NISP (n=6) | ISP<br>(n=125) | Active<br>Heme<br>Maligna<br>ncy<br>(n=10) | Inactive<br>Heme<br>Maligna<br>ncy<br>(n=21) | Solid<br>Maligna<br>ncy<br>(n=3) | Solid<br>Organ<br>Transpla<br>nt (n=24) | Autoim<br>mune<br>Disease<br>(n=38) | 1°<br>Immuno-<br>deficien<br>cy<br>(n=17) | Multiple<br>categori<br>es<br>(n=12) |
| On ISMT, % (n) | 0 (0) | 0 (0) | 71 (89) | 70 (7) | 43 (9) | 33 (1) | 100 (24) | 97 (37) | 6 (1) | 83 (10) |
| Male:female ratio | 0.2:1 | 0.2:1 | 0.8:1 | 1.5:1 | 1.3:1 | 2.0:1 | 2.0:1 | 0.5:1 | 0.4:1 | 0.5:1 |
| Age, median (IQR),<br>years | 54.0<br>(47.8-57.2) | 51.5 (42.5-58.2) | 58.0<br>(44.0-68.0) | 57.0<br>(38.5-69.5) | 65.0<br>(46.0-70.0) | 64.0<br>(63.5-73.5) | 50.0<br>(28.2-58.5) | 55.5<br>(45.8-64.0) | 63.0<br>(51.0-71.0) | 63.5<br>(54.2-71.0) |
| IgG days since<br>vaccine*, median (IQR) | 22.0<br>(16.2-29.2) | 34.0 (22.0-52.0) | 36.0<br>(28.2-49.8) | 27.0<br>(22.0-43.5) | 34.5<br>(30.0-49.0) | 27.0<br>(21.5-32.5) | 37.0<br>(27.0-46.5) | 37.0<br>(29.5-55.5) | 35.0<br>(31.0-42.0) | 44.0<br>(34.5-60.5) |
| Anti-S1 IgG, median<br>(IQR), OD ratio | 8.7 (7.7-9.7) | 9.8 (9.3-10.0) | 5.2 (0.3-8.6) | 1.5 (0.2-8.9) | 4.9 (0.1-8.7) | 8.1 (7.9-8.2) | 8.3 (0.4-9.1) | 3.3 (1.0-7.9) | 7.0 (4.3-7.8) | 1.7 (0.2-7.5) |
| Anti-S1 IgG, % positive<br>(n) | 100 (6/6) | 100 (5/5) | 69<br>(81/118) | 50 (5/10) | 55<br>(11/20) | 100 (2/2) | 70<br>(16/23) | 71<br>(25/35) | 88<br>(15/17) | 64 (7/11) |
| ACE2 Blocking %, median (IQR) | 95.5<br>(92.0-99.8) | 100.0<br>(100.0-100.0) | 31.5<br>(0.0-94.8) | 0.0 (0.0-87.0) | 26.0<br>(0.0-97.8) | 87.0<br>(82.5-91.5) | 90.0<br>(0.0-100.0) | 9.0 (0.0-42.5) | 36.0<br>(7.0-94.0) | 1.0 (0.0-66.0) |
| ACE2 Blocking, %<br>positive (n) | 100 (6/6) | 100 (5/5) | 53<br>(62/118) | 40 (4/10) | 50<br>(10/20) | 100 (2/2) | 65<br>(15/23) | 49<br>(17/35) | 59<br>(10/17) | 36 (4/11) |
| IGRA days since<br>vaccine^, median (IQR) | 22.0<br>(16.2-29.2) | 33.5 (24.8-47.5) | 36.5<br>(28.0-50.5) | 31.0<br>(22.0-46.2) | 39.0<br>(31.0-50.0) | 20.0<br>(18.0-29.5) | 36.5<br>(26.2-44.8) | 37.0<br>(28.0-57.0) | 35.0<br>(31.0-42.0) | 48.0<br>(34.8-73.0) |
| IFN-γ Response<br>median (IQR), IU/mL | 1.7 (1.0-4.0) | 4.7 (3.3-7.3) | 2.8 (0.3-8.8) | 9.9 (3.6-10.0) | 2.6 (1.2-6.0) | 9.3 (5.0-9.6) | 1.0 (0.1-3.2) | 3.9 (0.7-9.2) | 4.0 (1.4-8.9) | 0.7 (0.2-2.8) |
| IFN-γ Response, %<br>positive (n) | 100 (6/6) | 83 (5/6) | 73<br>(91/124) | 80 (8/10) | 76<br>(16/21) | 100 (3/3) | 54<br>(13/24) | 84<br>(31/37) | 82<br>(14/17) | 50 (6/12) |

Note: not all patients had both anti-S1 IgG and IGRA results available.
\*Number of days elapsed between primary vaccination and anti-S1 IgG testing.
^Number of days elapsed between primary vaccination and IGRA testing.

Immunosuppressive conditions were subcategorized by disease mechanism (eTables 3,4) and stratified into the following categories: hematologic malignancy with active disease at the time of initial vaccination or anytime between vaccination and testing (active heme malignancy), hematologic malignancy with complete response to therapy as per imaging or pathologic diagnosis (inactive heme malignancy), primary hematologic disease of anemia (primary anemia), solid non-hematologic malignancy with prior or active chemotherapy or radiation therapy (solid malignancy), solid organ transplant, autoimmune disease, and primary immunodeficiency. Patients without immunosuppressive diseases or history of ISMT use were included in the NISP (non-immunosuppressed patient) cohort. All other patients were included in the ISP (immunosuppressed patient) cohort. Patients with a documented history of SARS-CoV-2 infection or without vaccination, disease, or therapy documentation in the EMR were excluded from the analysis.

Immunocompetent healthcare worker (HCW) volunteers without known history of SARS-CoV-2 infection served as a reference cohort. Anti-S1 IgG and IGRA were collected between 14 and 25 days and then at approximately 5- and 9-months following primary vaccination with BNT162b2. Anti-S1 IgG and IGRA were collected between 8- and 34-days following receipt of a booster dose of the BNT162b2 vaccine in a subset of these HCWs.

### Statistical analysis

Statistical analyses and graphing were performed in Python version 3.8.5 using the packages pandas, matplotlib, seaborn, numpy, scipy, and statsmodels. Linear regression was performed by method of ordinary least squares. Unless otherwise indicated, Fisher exact test was used for all statistical comparisons, and α=.05. See eMethods for detailed descriptions.

We focused the analysis on anti-S1 IgG and IGRA positivity rates, rather than quantitative values, which decline over time (eFigure 1)^9,14^. The HCW cohort anti-S1 IgG values were used to establish a reference range for expected (i.e. immunocompetent) anti-S1 IgG levels over time after primary vaccination. A cutoff was established based on this reference range for “high” IgG, indicating expected IgG levels over time, and “low” IgG (eMethods). The same cutoff was not established for IGRA results due to the stochasticity observed with this assay.

**Figure 1.**
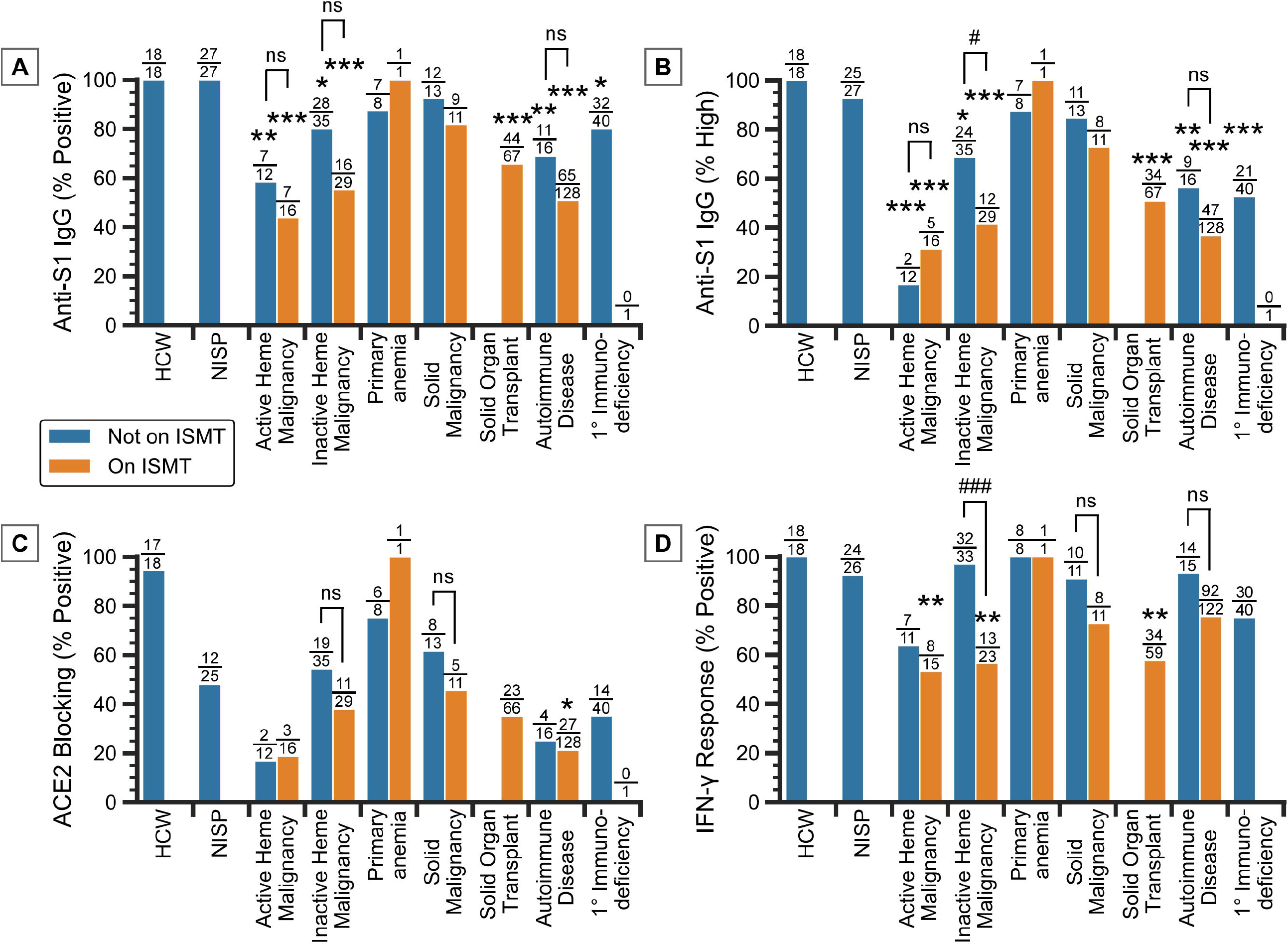
Positivity rates for the (A and B) IgG assay, (C) ACE2 blocking assay, and (D) IGRA in the HCW, NISP, and ISP cohorts categorized by disease and therapy status. Anti-S1 IgG positive cutoff is set at ≥1.1 (OD ratio). High anti-S1 IgG is determined by the lower bound of the 95% confidence interval (CI) calculated using equation 1 (eMethods). ACE2 blocking positive cutoff is set at ≥20%. IFN-γ response positive cutoff is set at ≥0.35 (IU/mL). Only patients with diseases belonging to a single category are shown. ISMT, immune-suppress/modulatory therapy; ns, not significant; ^#^P<.05, ^##^P<.01, ^###^P<.001 for pairwise comparisons as indicated; *P<.05, **P<.01, ***P<.001 compared to NISP, Fisher exact test.

## RESULTS

A total of 496 patients were included in this study. Cohort sample size and assay result availability are presented in eTable 5.

### Vaccine response after primary vaccination

Demographic information and assay results from 18 HCWs, 28 NISPs, and 427 ISPs following primary vaccination are displayed in Table 1 (see also eTables 6,7). The difference in anti-S1 IgG (62%) and IGRA (71%) positivity rates in ISPs was statistically significant (*P*=.009). In 381 ISPs that had both anti-S1 IgG and IGRA results available, 51% were positive for both (196), 18% were negative for both (67), 20% were IGRA positive only (75), and 11% were IgG positive only (43).

### Relative contributions of immunosuppressive diseases and ISMTs to impaired vaccine response

For ISPs with only a single disease category, the anti-S1 IgG and IGRA positivity rates were generally lower for those on ISMT than those not on ISMT, albeit this was only statistically significant for patients with inactive hematological malignancy (Figure 1). Overall, ACE2 blocking results showed similar positivity rates as anti-S1 IgG (Figure 1, eFigure 2).

**Figure 2.**
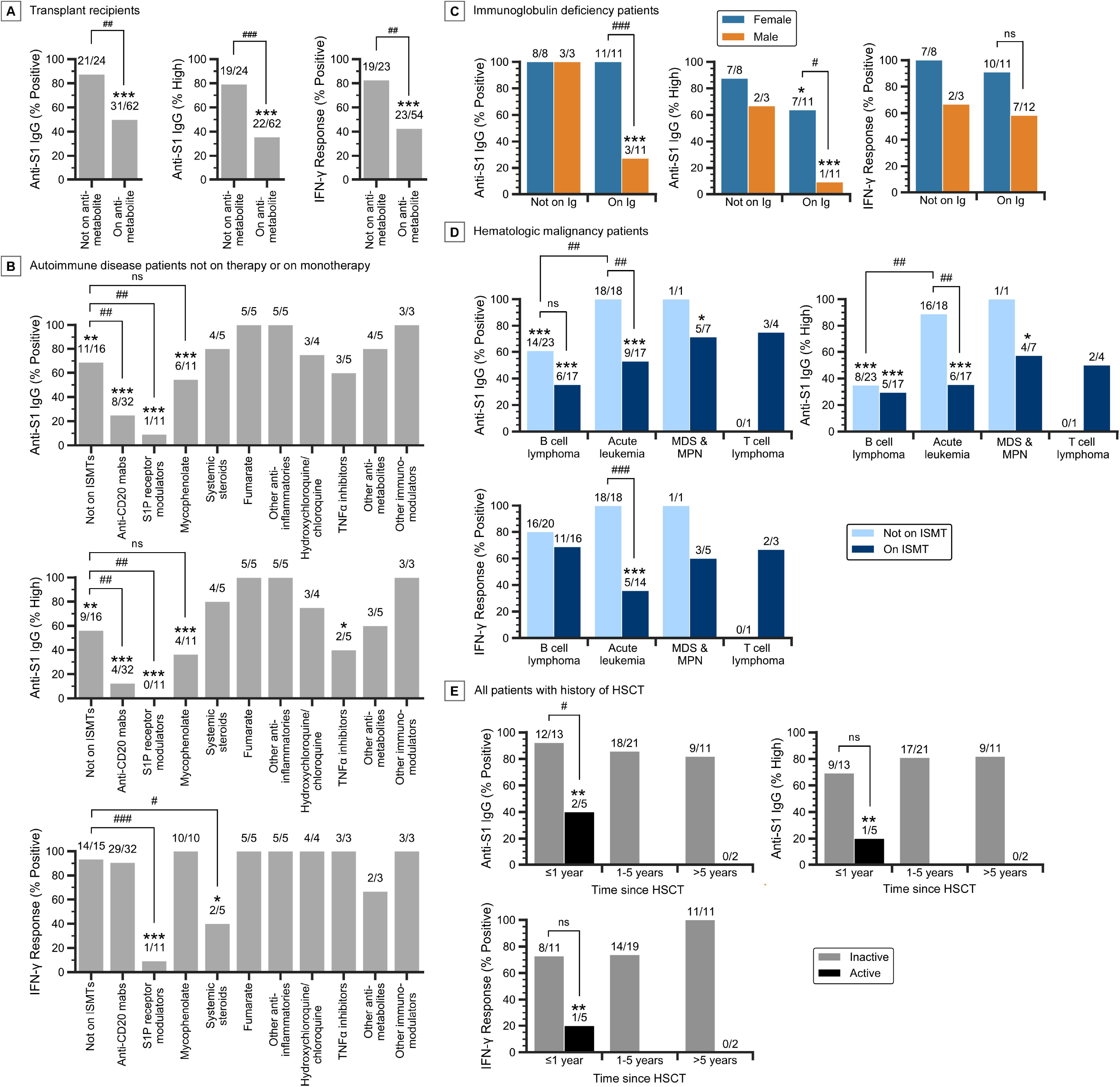
Subgroup analysis of immunosuppressed patients following primary vaccination. A) Vaccine response rates in transplant recipients not on antimetabolites and those on antimetabolites (61 on mycophenolate, 1 on azathioprine). Only transplant recipients on a calcineurin inhibitor, mTOR inhibitor, mycophenolate, or other antimetabolites are plotted. B) Vaccine response rates of autoimmune disease patients, without other immunosuppressive conditions, on monotherapy with various ISMTs. Only ISMTs that apply to three or more patients are plotted. C) Vaccine response rates of immunoglobulin deficiency patients, without other immunosuppressive conditions, stratified by immunoglobulin use and gender. D) Vaccine response rates of patients with active and inactive heme malignancy, without other immunosuppressive conditions, stratified by disease subcategory and therapy status. Only disease subcategories that apply to three or more patients are plotted. E) Vaccine response rates of all patients with a history of HSCT, stratified by time between HSCT and vaccination, and activity of the malignancy. Patients with inactive disease include nine patients with primary anemia and one patient with germ cell tumor. ns, not significant; ^#^*P*<.05, ^##^*P*<.01, ^###^*P*<.001 for pairwise comparisons as indicated; \**P*<.05, \*\**P*<.01, \*\*\**P*<.001 compared to NISP, Fisher exact test.

For patients not on ISMTs, two categories had high rates of humoral and cellular response, comparable to those of NISPs: primary anemia and solid malignancy. Primary anemia patients not on ISMT were 88% IgG positive (7/8), 88% IgG high (7/8), and 100% IGRA positive (8/8). Solid malignancy patients were 92% IgG positive (12/13), 85% IgG high (11/13), and 91% IGRA positive (10/11). Notably, solid malignancy patients on ISMT also had relatively high response rates compared to NISPs, at 82% IgG positive (9/11, *P*=.08), 73% IgG high (8/11, *P*=.13), and 73% IGRA positive (8/11, *P*=.14).

Two categories of patients not on ISMTs had low rates of humoral response compared to NISPs but high rates of cellular response: autoimmune disease and inactive hematologic malignancy. Autoimmune disease patients not on ISMT were 69% IgG positive (11/16, *P*=.005), 56% IgG high (9/16, *P*=.008), and 93% IGRA positive (14/15). Inactive hematologic malignancy patients were 80% IgG positive (28/35, *P*=.02), 69% IgG high (24/35, *P*=.03), and 97% IGRA positive (32/33).

Primary immunodeficiency and active hematologic malignancy patients not on ISMT had low rates of both humoral and cellular response; however the rates of cellular response were not significantly lower than that of NISPs. Primary immunodeficiency patients were 80% IgG positive (32/40, *P*=.02), 53% IgG high (21/40, *P*<.001), and 75% IGRA positive (30/40, *P*=.11). Active hematologic malignancy patients were 58% IgG positive (7/12, *P*=.001), 17% IgG high (2/12, *P*<.001), and 64% IGRA positive (7/11, *P*=.05).

### Immunosuppressive factors associated with poor vaccine response

Results of a systematic screen of immunosuppressive factors associated with low and high rates of humoral and cellular response are shown in eTables 8-11 and eFigures 3,4. Notable factors associated with a low rate of humoral and cellular response include S1P receptor modulators (IgG: n=11, *P*<.001; IGRA: n=11, *P*<.001), mycophenolate (IgG: n=78, *P*=.002; IGRA: n=69, *P*<.001), and systemic steroids (IgG: n=103, *P*=.002; IGRA: n=93, *P*<.001). Anti-CD20 mAbs (n=48, *P*<.001), autoimmune disease (n=173, *P*<.001), active heme malignancy (n=34, *P*=.002), B cell lymphoma (n=55, *P*=.004), and immunoglobulins (n=50, *P*=.01) were associated with a low rate of humoral response specifically, while calcineurin inhibitors (n=89, *P*<.001), solid organ transplant (n=79, *P*<.001), and antimitotics (n=13, *P*<.009) were associated with a low rate of cellular response specifically. Conversely, liver transplant (n=15, *P*<.001) and solid malignancy (n=34, *P*=.001) were associated with a high rate of humoral response, while anti-CD20 mAbs, specifically ocrelizumab (n=24, *P*=,008) were associated with a high rate of cellular response.

**Figure 3.**
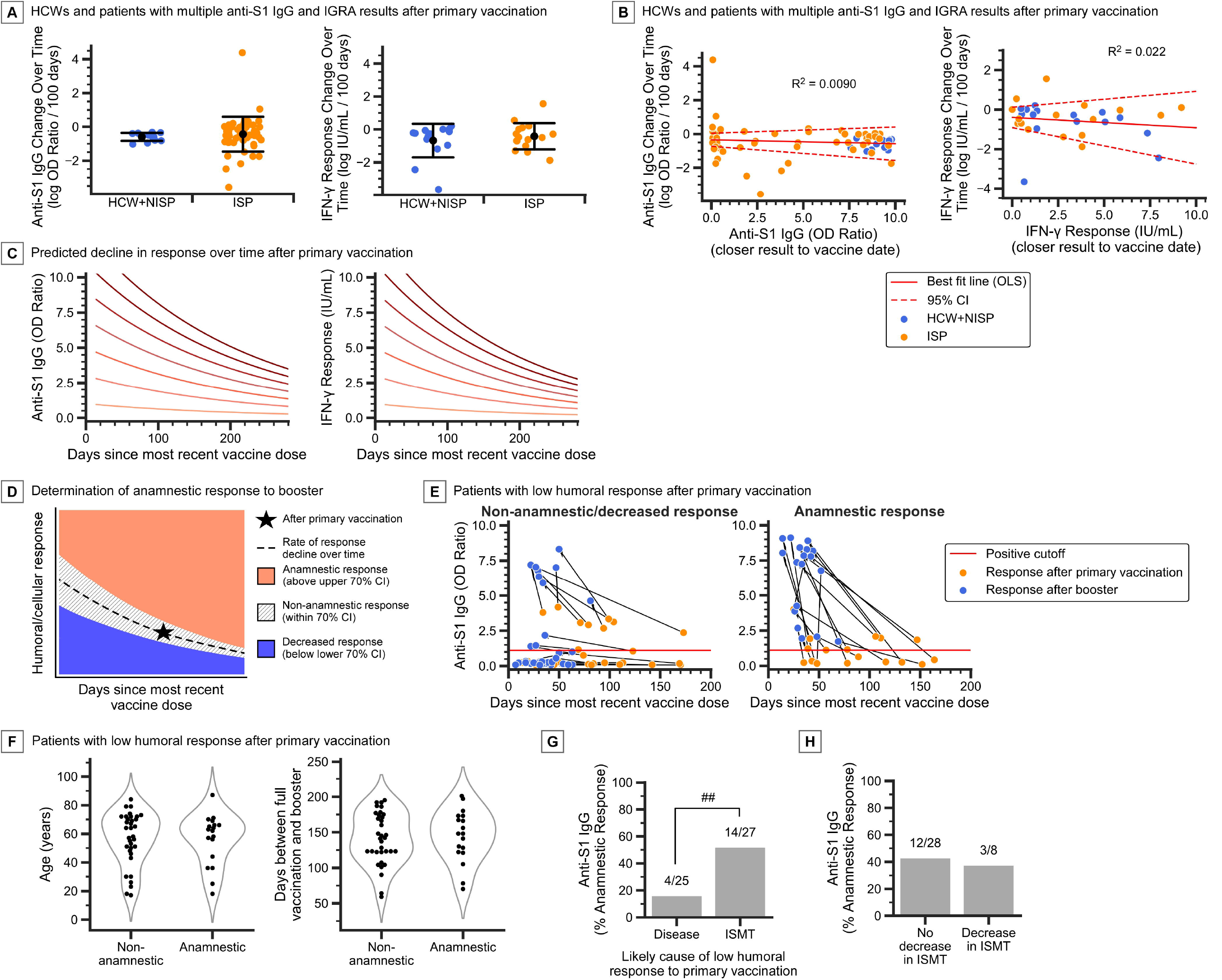
Change in vaccine response rates over time and after booster vaccination. A) Comparison of the change in natural log transformed anti-S1 IgG and IGRA values over time in HCW, NISP, and ISP cohorts. Error bars, mean ± standard deviation. B) Linear model fitted to the change in natural log transformed vaccine response over time versus the initial vaccine response (using the value of the assay performed closest to the vaccination date). Dotted lines denote the 95% confidence interval (CI) of the linear model mean and intercept. Note the 95% CI for slope includes 0 in both cases. C) Predicted decline in vaccine response over time using equations 2 and 3 (eMethods), at various starting (i.e. theoretical peak) response values after primary vaccination. D) Determination of anamnestic booster response given primary vaccination (primary dose) response (Star), based on the expected rate of change in response over time after primary vaccination (dashed line), with confidence intervals (CI) as calculated using equations 2 and 3 (eMethods). Booster responses that fall above the upper bound of the 70% CI (pink region) are determined to be anamnestic. E) ISP (n = 52) cohort with low anti-S1 IgG after primary vaccination separated into non-anamnestic/decreased booster response (n = 34) and anamnestic booster response (n = 18), showing both the response after primary vaccination (orange dots) and after booster (blue dots). F) Distribution of age and days between primary vaccination and booster in non-anamnestic and anamnestic booster response patients. G) Anamnestic booster response rates in patients stratified by the likely primary cause of low humoral (anti-S1 IgG) response after primary vaccination (disease: heme malignancy and primary immunodeficiency patients, ISMT: solid malignancy, solid organ transplant, and autoimmune disease patients) H) Anamnestic booster response rates in patients stratified by whether there was a decrease in ISMT dosage around the time of booster vaccination. Only patients on ISMT during the primary doses are included. Panels A and B: n=12 HCWs, 2 NISPs, 52 ISPs (IgG) and n=15 HCWs, 1 NISP, 15 ISPs (IGRA). Panels F-H: only patients with low anti-S1 IgG after primary vaccination are included. ns, not significant; ^#^*P*<.05, ^##^*P*<.01, ^###^*P*<.001 for pairwise comparisons as indicated, Fisher exact test.

Results of multivariable linear regression to identify individual effects of immunosuppressive factors and certain demographic factors on humoral and cellular response are presented in eTable 12,13.

### Subgroup analysis to verify findings of immunosuppressive screen and regression modeling

86 solid organ transplant recipients were on ISMT regimens consisting of tacrolimus (n=75), mycophenolate (61), systemic steroids (52), sirolimus (5), cyclosporine (5), everolimus (3), or azathioprine (1). Of these patients, the addition of antimetabolites to a drug regimen consisting of calcineurin or mTOR inhibitors, with or without systemic steroids, is associated with low humoral and cellular response, as patients not receiving antimetabolites were 88% IgG positive (21/24), 79% IgG high (19/24), and 83% IGRA positive (19/23), while those on antimetabolites (n=61 mycophenolate, n=1 azathioprine) were 50% IgG positive (31/62, *P*=.001), 35% IgG high (22/62, *P*<.001), and 43% IGRA positive (23/54, *P*=.001) (Figure 2A). Further stratifying transplant recipients by age, time between transplantation and vaccination, and organ type showed that age and the amount of time between transplant and vaccination may have small effects on immune response (eResults, eFigures 5,6).

Vaccine responses of 110 autoimmune disease patients, not affected by diseases of any other category and either on monotherapy or not actively on ISMT, are shown in Figure 2B. Compared to autoimmune patients not on ISMT, patients on S1P receptor modulators had low humoral response rates, at 9% IgG positive (1/11, *P*=.004) and 0% IgG high (0/11, *P*=.003). Likewise, patients on anti-CD20 mAbs were 25% IgG positive (8/32, *P*=.005) and 12.5% IgG high (4/32, *P*=.004). Importantly, while patients on S1P receptor modulators were 9% IGRA positive (1/11, *P*<.001), those on anti-CD20 mAbs were 91% IGRA positive (29/32). Compared to autoimmune disease patients not on ISMT, those on systemic steroids had low cellular response, at 40% IGRA positive (2/5, *P*=.03), while those on fumarate (5/5) and other anti-inflammatories (5/5) had 100% positive humoral and cellular responses. Autoimmune patients on mycophenolate had lower rates of humoral response compared to NISPs, at 55% IgG positive (6/11, *P*<.001) and 36% IgG high (4/11, *P*<.001), but were 100% IGRA positive (10/10). Notably, the vaccine response rates in patients on mycophenolate are comparable to those of autoimmune disease patients not on ISMTs.

Analysis of 41 patients with immunoglobulin deficiency, without other immunosuppressive diseases and not on any ISMT, was underpowered to definitively identify a difference in vaccine response due to immunoglobulin therapy. However, while female patients on immunoglobulin therapy were 100% IgG positive (11/11), 64% IgG high (7/11), and 91% IGRA positive (10/11), male patients had much lower rates especially with humoral response, at 27% IgG positive (3/11, *P*=.001), 9% IgG high (1/11, *P*=.02), and 58% IGRA positive (7/12, *P*=.15) (Figure 2C, eFigure 7).

Analysis of 94 patients with active and inactive hematologic malignancy showed that in patients not on ISMT, those with B cell lymphoma were 61% IgG positive (14/23) and 35% IgG high (8/23), while those with acute leukemia (12 B-ALL, 5 AML, and 1 MPAL) had much higher rates, at 100% IgG positive (18/18, *P*=.002) and 89% IgG high (16/18, *P*<.001) (Figure 2D, eFigure 8). The cellular response rates in these patients were both high, however, with B cell lymphoma patients at 80% IGRA positive (16/20) and acute leukemia patients at 100% IGRA positive (18/18, *P*=.11).

While acute leukemia patients not on ISMTs had high rates of humoral and cellular response, those on ISMTs (9 B-ALL, 5 AML, and 3 T-ALL) had much lower rates, at 53% IgG positive (9/17, *P*=.001), 35% IgG high (6/17, *P*=.002), and 36% IGRA positive (5/14, *P*<.001). Interestingly, B cell lymphoma patients on ISMTs, at 35% IgG positive (6/17, *P*=.20), 29% IgG high (5/17, *P*>.99), and 69% IGRA positive (11/16, *P*=.47), did not have lower response rates than B cell lymphoma patients not on ISMTs (Figure 2D).

For 52 patients with history of HSCT, which included 42 patients with a hematologic malignancy, nine with primary anemia, and one with germ cell tumor, analysis showed that HSCT patients without active hematologic malignancy had high rates of humoral and cellular response, at 87% IgG positive (39/45), 78% IgG high (35/45), and 80% IGRA positive (33/41). By comparison, HSCT patients with active heme malignancy (either newly diagnosed or relapsed/residual disease) had much lower rates of response, at 29% IgG positive (2/7, *P*=.003), 14% IgG high (1/7, *P*=.002), and 14% IGRA positive (1/7, *P*=.001).

Patients who received HSCT within one year prior to vaccination, without active hematologic malignancy, were 92% IgG positive (12/13), 69% IgG high (9/13), 73% IGRA positive (8/11) (Figure 2E). These response rates were comparable to those who received HSCT greater than one year prior to vaccination, which were 84% IgG positive (27/32), 81% IgG high (26/32), and 83% IGRA positive (25/30).

### Analysis of the rate of decline in quantitative vaccine response over time after primary vaccination

Patients with multiple anti-S1 IgG and IGRA results after primary vaccination showed that the average rates of decline for both humoral and cellular response in ISPs were comparable to those of HCWs and NISPs (Figure 3A, eFigure 9). The average rate of change of log transformed anti-S1 IgG OD ratio is - 0.602 per 100 days in HCWs and NISPs (SD=0.264), and -0.434 per 100 days in ISPs (SD=1.05, *P*=.56, two-tailed t test). The average rate of change of log IFN-γ response (IU/mL) is -0.682 per 100 days in HCWs and NISPs (SD=1.02), and -0.411 per 100 days in ISPs (SD=0.849, *P*=.43, two-tailed t test).

Notably, the standard deviation in the rate of humoral response change over time was 4-fold higher in ISPs than in HCWs and NISPs combined. For cellular response, the standard deviation of the rate of change is much more comparable between ISPs and HCWs plus NISPs, with a 0.8-fold difference.

Importantly, the rate of change in vaccine response over time did not correlate with the initial anti-S1 IgG (best fit line slope=-2.31E-4, 95% CI -8.35E-4 to 3.74E-4, *P*=.45, two-tailed Pearson correlation) or IGRA (best fit line slope=-5.23E-4, 95% CI -1.85E-3 to 8.07E-4, *P*=.43, two-tailed Pearson correlation) values (Figure 3B). For anti-S1 IgG, the relationship between rate of change and initial value displays heteroscedasticity, where the variance in the rate of change was higher towards the low end of the initial value. After investigation, we attributed this heteroscedasticity mainly to the imprecision of the anti-S1 IgG assay, which particularly at low values, become magnified by log transformation (eResults).

### Vaccine response after booster vaccination

Demographic information and assay results from 6 immunocompetent HCWs, 6 NISPs, and 125 ISPs after booster vaccination are displayed in Table 2 (see also eTables 14,15). The difference in anti-S1 IgG (69%) and IGRA (73%) positivity rates in ISPs was not statistically significant (*P*=.48). In 117 ISPs that had both anti-S1 IgG and IGRA results available, 55% were positive for both (64), 14% were negative for both (16), 18% were IGRA positive only (21), and 14% were IgG positive only (16).

In the paired booster cohort of 6 HCWs, 1 NISP, and 84 ISPs with anti-S1 IgG or IGRA results collected before and after the booster dose, testing occurred a median of 89 days after primary vaccination, and 36 days after booster. To correct for this collection time difference, we applied the average rate of vaccine response decline over time after the primary dose (Figure 3C) to predict the expected non-anamnestic booster responses (where the response after primary and booster doses are the same). This allowed us to determine, with statistical confidence, patients who had anamnestic booster responses (Figure 3D, eFigure 10, eMethods, eResults). Of 40 ISPs with both paired anti-S1 IgG and paired IGRA results available, 10 had an anamnestic humoral booster response and two had an anamnestic cellular booster response, a statistically significant difference (*P*=.03). Notably, 35% (18/52) ISPs with low humoral response after primary vaccination had an anamnestic booster response (Figure 3E). We focused the rest of the analysis on humoral booster response, and specifically on patients with low humoral response after the primary doses.

Patient age and duration between primary vaccination and booster doses were not associated with booster response (Figure 3F). Patients with immune defects due to disease and not necessarily ISMT (hematologic malignancy and primary immunodeficiency patients) had low rates of anamnestic response of 16% (4/25), while patients with immunodeficiency due to ISMT use (solid malignancy, solid organ transplant, and autoimmune disease patients) had higher rates of anamnestic response of 52% (14/27, *P*=.009) (Figure 3G).

Following boosters, 50% of patients on anti-CD20 mAbs had an anamnestic response (4/8), 0% for S1P receptor modulators (0/2), 56% for mycophenolate (5/9), and 50% for systemic steroids (5/10). 20% of patients with acute leukemia had an anamnestic response (1/5), 18% for B cell lymphoma (2/11), 33% for plasma cell disease (1/3), and 0% for CVID (0/4) (eFigure 11).

Eight patients had their ISMT dosage temporarily decreased by their provider to try to elicit an anamnestic booster response. Three of these patients had an anamnestic response (38%), a rate comparable to patients who did not have their ISMT altered (43%, 12/28) (Figure 3H, eResults).

## DISCUSSION

We identified patient factors affecting immune response to primary and booster SARS-CoV-2 vaccines in a large heterogeneous cohort of immunosuppressed patients. We found more immunosuppressive factors associated with low humoral response than low cellular response, corresponding with the lower rate of anti-S1 IgG positivity (62%) compared to IGRA (71%) in immunosuppressed patients. In concordance with other studies^15–23^, we found that anti-CD20 mAb and S1P receptor modulator use are associated with decreased humoral response. Unlike anti-CD20 mAb use, S1P receptor modulator use is also associated with low rates of cellular response. We showed that antimetabolite/mycophenolate use is the strongest predictor of decreased humoral and cellular response in transplant recipients, in concordance with other studies^4^. In autoimmune patients, however, mycophenolate monotherapy was not definitively associated with decreased humoral or cellular response. While monotherapy with either a calcineurin/mTOR inhibitor or mycophenolate was not associated with decreased vaccine response, combination therapy was. HSCT patients without active hematologic malignancy had relatively high rates of humoral and cellular response, even if vaccinated less than one year from transplantation. This is concordant with findings from one study^24^, but not another^25^. Notably, results from the latter study may be confounded by recurrent/residual disease after HSCT.

Hematological malignancies were associated with lower vaccine response than solid malignancies, consistent with other reports^15,16,20,22,26,27^. Interestingly, patients with a history of B cell lymphomas or plasma cell diseases, not on ISMT, had low rates of humoral response but high rates of cellular response, while those with a history of acute leukemias, such as B lymphoblastic leukemia, had high rates of both humoral and cellular response. Primary hypogammaglobulinemia disorders were associated with decreased humoral response, particularly in male patients. It has been shown that men typically produce lower antibody responses to non-SARS-CoV-2 vaccines^28^, an effect likely amplified in hypogammaglobulinemia.

Importantly, we did not find evidence that the rate of decline in humoral and cellular response over time differ between non-immunosuppressed individuals and ISPs. This allowed us to predict the expected non-anamnestic and anamnestic booster responses for each patient.

We find no strong evidence that boosters improve cellular response in ISPs, in concordance with a previous report in cancer patients^29^, but contradicting other studies^23,30^. The rate of anamnestic humoral response after booster in patients with poor humoral response after primary vaccination was 35%.

Interestingly, patients with poor response to primary vaccination due at least in part to the immunosuppressive effect of their primary disease were unlikely to have an anamnestic booster response. Meanwhile, patients with poor response to primary vaccination due to therapy were more likely to have an anamnestic booster response. This suggests that the immunosuppressive effects of ISMTs, such as anti-CD20 mAbs, can be overcome with booster immunization. Finally, although the analysis may be underpowered, we find no evidence that temporary ISMT dose reduction leads to a higher rate of anamnestic booster response.

Although this study was strengthened by evaluating vaccine responses in a large heterogeneous cohort of immunosuppressed patients, limitations include the small sample size of certain patient disease and ISMT categories, and a lack of information on drug dosages and history of prior therapy. Additionally, there were no pediatric patients included in NISP cohort.

As the COVID-19 pandemic ensues and new SARS-CoV-2 variants arise, our findings provide an evidence-based framework for clinicians to determine optimal vaccination strategy in immunosuppressed patients. While numerous studies have examined humoral response to SARS-CoV-2 vaccination in particular immunosuppressed subgroups, relatively few have examined concurrent cellular response. This study shows that immunosuppressive conditions differentially impact humoral and cellular responses to SARS-CoV-2 vaccines, with 20% of patients only developing a cellular response following initial vaccination. The importance of cellular response in anti-SARS-CoV-2 immunity is supported by several reports^8,31–35^ and a recent study showed that emerging SARS-CoV-2 variants that escape humoral immunity in vaccinees may not escape cellular immunity^36^. Humoral and cellular responses, therefore, provide complementary protection against SARS-CoV-2 in immunosuppressed patients. This emphasizes the importance of monitoring both humoral and cellular responses to vaccination, especially in immunosuppressed patients, and the utility of performing cellular immunity testing in the clinical laboratory.

## Data Availability

De-identified raw data used in the study is available upon request.

## ABBREVIATIONS

CLL: chronic lymphocytic leukemia
CVID: common variable immunodeficiency
HCW: healthcare worker
HSCT: hematopoietic stem cell transplant
Ig: immunoglobulins
IgG assay: anti-S1 IgG enzyme-linked immunosorbent assay (Euroimmun)
IGRA: interferon-γ (INF-γ) release assay (Stanford)
ISMT: immune-suppressive/modulatory therapy
ISP: immunosuppressed patient
mAbs: monoclonal antibodies
MBL: monoclonal B cell lymphocytosis
MDS: myelodysplastic syndrome
MGUS: monoclonal gammopathy of unknown significance
MPN: myeloproliferative neoplasm
MS: multiple sclerosis
NISP: non-immunosuppressed patient
NMO: neuromyelitis optica
S1P: sphingosine 1-phosphate
SLE: systemic lupus erythematosus
SLL: small lymphocytic lymphoma

## DATA AVAILABILITY

De-identified raw data used in the study is available upon request.

## ACKNOWLEDGEMENTS

We thank all study participants, providers and nursing staff caring for immunocompromised patients evaluated in this study.

