## Supplemental materials for "Cell-mediated and humoral immune response to SARS-CoV-2 vaccination and booster dose in immunosuppressed patients"

### eMethods

#### Assay Design

The SARS-CoV-2 interferon gamma (IFN-γ) release assay (IGRA) is a laboratory-developed *in vitro* blood diagnostic used to measure IFN-γ released by SARS-CoV-2 antigen-specific T cells following overnight stimulation with pathogen-specific peptides, as previously described[1]. Humoral response was evaluated by commercial anti-SARS-CoV-2 spike S1 domain IgG (anti-S1 IgG) ELISA (Euroimmun; Lübeck, Germany). Plasma samples with positive anti-S1 IgG were reflexed to a laboratory-developed assay to measure the percentage of antibodies blocking the interaction between purified RBD protein and recombinant human angiotensin-converting enzyme 2 (ACE2) receptor in an ACE2 competition ELISA, performed as a surrogate measurement of neutralizing antibodies (ACE2 blocking activity)[2]. Patients with negative anti-S1 IgG were considered to have 0% blocking activity. Laboratory methods and assay details are further described below.

#### Interferon Gamma Release Assay (IGRA)

SARS-CoV-2 IGRA was performed in the Stanford Health Care Clinical Microbiology Laboratory as described previously[1]. Briefly, freshly collected blood in lithium heparin tube was transferred to BD vacutainer no additive tubes at 1mL per tube. One tube was left unstimulated (nil), one tube was stimulated with SARS-CoV-2 antigen peptide megapool (Miltenyi Biotec, Bergisch Gladbach, Germany) at 2.2 mmol/mL, and one tube was stimulated with Phytohemagglutinin PHA-P Mitogen (Sigma, St. Louis, MO) at 50µL/mL. The blood samples were mixed gently and incubated at 37 °C for 20 to 24 h. The interferon-γ concentration was measured in the plasma portion with an automated enzyme-linked immunosorbent assay (ELISA) instrument (DSX; Dynex Technologies, Chantilly, VA) using the QuantiFERON-TB ELISA kit (Qiagen, Germantown, MD). Interferon-γ response was defined as positive if antigen-nil ≥0.35 IU/mL; negative if antigen-nil <0.35 and mitogen-nil ≥0.5 IU/mL, and indeterminate if nil >8 IU/mL or antigen-nil <0.35 and mitogen-nil <0.5 IU/mL. All values higher than 10 IU/mL were adjusted to 10 IU/mL to reflect the linear range of assay.

#### Enzyme-linked immunosorbent assay (ELISA)

SARS-CoV-2 Spike S1 IgG serology was performed by semi-quantitative ELISA (EUROIMMUN, Lübeck, Germany) according to the manufacturer's instructions. Briefly, 1:100 diluted serum samples from vaccinees were added to the antigen pre-coated microplate followed by the addition of anti-human IgG conjugate, chromogenic substrate and stopping solution. The optical density (OD) was measured at 450nm. Results were evaluated by calculating the ratio of the optical density (OD) of the sample over the OD of a calibrator provided by the manufacturer. Manufacturer assay cutoff values were defined as: <1.1 negative, ≥1.1 positive. All values higher than OD 10 were adjusted to 10 to reflect the linear range of assay.

#### ACE2 RBD blocking activity assay (ELISA)

Anti-RBD IgG present in plasma samples sufficient to block binding between purified RBD protein and recombinant human angiotensin-converting enzyme 2 (ACE2) receptor were measured using a laboratory-developed competition ELISA, performed on the ESP 600 instrument as previously described[2]. In brief, RBD-coated plates are incubated with plasma at a 1:10 dilution, recombinant ACE2 joined to a mouse IgG2a Fc (ACE2-mFc) is added at 0.5 µg/mL, and secondary detection is performed using horseradish peroxidase conjugated goat anti-mouse IgG at a 1:10,000 dilution. Samples are run in duplicate, and average OD at 450 nm is used to calculate the sample-to-negative calibrator ratio. Blocking activity is reported as a percentage as follows: (1-ratio) x 100, with higher percentages corresponding to lower levels of RBD-ACE2 binding. Cutoff value for positive ACE2 blocking was set at ≥20%.

#### Immunosuppressive factor screen

Fisher exact test was used to detect immunosuppressive factors (therapies and diseases) associated with a specified outcome, which were then ranked by P value. NISP and ISP cohorts are included. Immunosuppressive factors are all categorical variables: immune lineage affected by primary disease (myeloid, B, or T), disease categories, disease subcategories, therapy categories, and on any ISMT. Multiple comparisons correction was applied to determine statistically significant P values using the Benjamini-Hochberg method, with a false discovery rate of 10%.

#### Linear regression and dominance analysis

Linear regression model fitting was performed in Python using the package statsmodels, via the method of ordinary least squares. Categorical variables were encoded as dummy variables 0 and 1, where 1 indicates patient is affected by the variable. Gender was encoded as 0 for female and 1 for male. Categorical variables that included n<3 0s or n<3 1s were excluded.

Dominance analysis to determine relative importance of individual independent variables in regression modeling was performed in Python using the library Dominance-Analysis (https://github.com/dominance-analysis/dominance-analysis), with default settings.

**Determination of lower than expected humoral response (low anti-S1 IgG)**

The time between vaccination and testing is a confounding variable in the dataset. However, we chose not to adjust assay quantitative results for this confounder. There are three reasons for this decision: 1) the imprecision of the assays, 2) the limited linear range of the assays, and 3) the stochasticity in the IGRA results. To control for the decline in anti-S1 IgG over time after vaccination, we established a reference interval for expected responses in immunocompetent individuals over time utilizing the HCW results collected at different timepoints after primary vaccination. A linear model correlating time since vaccine administration and natural log transformed anti-S1 IgG and IGRA values was fitted by the ordinary least square method (eFigure 1A). For this modeling, only the 17 HCWs with repeat anti-S1 IgG assays were included, and only two datapoints were included for each HCW (the two furthest apart in time of collection). A 95% confidence interval (CI) around the best fit line was calculated using the residuals from all HCW datapoints. NISP and ISP anti-S1 IgG values that fell above the upper bound of the 95% CI represent optimal anti-S1 IgG response to vaccination, while those that fell below the lower bound are suboptimal (eFigure 1B). The former values are considered “high anti-S1 IgG” and the latter “low anti-S1 IgG” (eFigure 1C). The same analysis was performed for IFN-γ response. IGRA values resulted as 0 were converted to 0.01 to perform natural log transformation. Due to the inter-individual stochasticity of the IGRA results, we did not classify them as high and low. The lower bound of the 95% CI around the linear model fitted to HCW results is given by the following equation, where *t* is the time since the most recent vaccine dose, in days:

Equation 1:

$$IgG_{lower\ bound} = e^{-0.004676t+1.724}$$

**Determination of vaccine response dynamics and anamnestic booster response**

We analyzed patients with multiple specimens collected for anti-S1 IgG assay and IGRA at different timepoints after primary vaccination, prior to booster, to determine the average rate of change in vaccine response over time. Only two assay results were used per patient in those with more than two results available (the two results furthest apart in collection time were used). Patients with anti-S1 IgG or IFN-γ response values above the linear range of the assays or with repeat collections less than 30 days apart were excluded from this analysis to improve accuracy and precision. This resulted in a sample size of 66 subjects (12 HCWs, 2 NISPs, 52 ISPs) with two IgG results and 31 subjects (15 HCWs, 1 NISP, 15 ISPs) with two IGRA results.

Given an individual’s anti-S1 IgG and IFN-γ response values at one timepoint after vaccine administration (after peak response has been reached, i.e. after ~14 days), we predicted the response values at any other timepoint using the following equations:

Equation 2:

$$IgG_{predicted} = e^{-0.004693t\ +\ b\ \pm\ \varepsilon}$$

Equation 3:

$$IGRA_{predicted} = e^{-0.005511t\ +\ b\ \pm\ \varepsilon}$$

Where -0.004693 is the average rate of decline in natural log transformed anti-S1 IgG values after primary vaccination in all individuals in the dataset with multiple IgG assays after primary vaccination; -0.005511 is the average rate of decline in natural log transformed IGRA values after primary vaccination in all individuals with multiple IGRA assays after primary vaccination; *t* is the time elapsed since primary vaccination, in days; *b* represents the theoretical IgG or IGRA peak (at day 0), which can be calculated with the given *t* and IgG/IGRA values; *ε* is the error term, which is provided by ($\bar{x}_{error}+s_{error}*z$): mean plus standard deviation of the prediction error multiplied by a chosen z-score (z = 1.036 for a 70% confidence interval, and z = 1.960 for a 95% confidence interval). Prediction error is given by the difference between natural log transformed observed and expected values when the equations are used to predict the repeat IgG/IGRA result when given the initial result ($\bar{x}_{error}$ = 0.06188, $s_{error}$ = 0.6610 for anti-S1 IgG, and $\bar{x}_{error}$ = -0.02243, $s_{error}$ = 1.360 for IGRA).

The same equations were then used to predict booster vaccination responses, given patients' responses to primary dose. We considered actual booster responses that fell within the 70% CI of the predicted response to be non-anamnestic, those that fell below the lower bound of the 70% CI to be decreased response, and those that fell above the upper bound of the 70% CI to be anamnestic response (Figure 3D). Patients identified as having non-anamnestic booster responses were excluded from the analysis if their post-booster IgG or IGRA values are above the linear range of the assay, as we cannot determine their true booster response. This applied to 7 patients in the humoral response analysis, and 11 patients in the cellular response analysis.

### eResults

#### Factors not associated with immunosuppression that affect anti-S1 IgG and IGRA results

As vaccine response has been shown to decline logarithmically over time, we suspected that time between vaccination and testing may be a major confounding factor in our dataset[3,4]. Prior studies have also demonstrated that age and possibly gender may also affect vaccine response[3,5,6]. To investigate how these factors independently affect anti-S1 IgG and IGRA values in our patient population, we performed multivariable linear regression with dominance analysis with the NISP cohort.

For humoral response, the combination of time since vaccine administration, age, and gender predicts log transformed anti-S1 IgG value with an $R^2$ = 0.497. Time since vaccine administration is the most predictive factor, with an incremental $R^2$ = 0.424, followed by age (0.050), and then gender (0.023). For cellular response, the combination of these three factors poorly predicts log transformed IGRA value, with an $R^2$ = 0.159. Time since vaccine administration is the most predictive factor (incremental $R^2$ = 0.100), followed by age (0.051), and then gender (0.008). Even in the HCW cohort, IGRA results varied dramatically (eFigure 2C), demonstrating an unexplained inter-individual stochasticity in cellular response as measured by IGRA. As ACE2 blocking percent correlates with anti-S1 IgG titers (eFigure 1D)[2], we did not perform a similar analysis. Given the effect of time since vaccine administration on anti-S1 IgG assay results and the stochasticity of IGRA results, we analyzed the rates of positivity of the two tests, as well as that of ACE2 blocking, rather than the values.

To control for the effect of time since vaccine administration on IgG assay results, we established a reference range based on the change in anti-S1 IgG over time in the HCW cohort (eMethods). Nearly all NISPs (25/27) fell into the "high IgG" category (eFigure 1). We then screened for immunosuppressive factors that are enriched in patients with positive but low anti-S1 IgG relative to patients with positive and high anti-S1 IgG. The top 10 immunosuppressive factors enriched in patients with low positive anti-S1 IgG include active therapy with mycophenolate or immunoglobulins, and having plasma cell disease, an active heme malignancy, marginal zone lymphoma, or monoclonal gammopathy of unknown significance. All of these factors have either been shown previously to negatively affect humoral vaccine response or B cell function[7–10]. Meanwhile, among the top 10 factors enriched in patients with high positive anti-S1 IgG include having primary anemia and not receiving IMST. These findings suggest that comparing high versus low anti-S1 IgG results is likely more sensitive for detecting a defect in humoral vaccine response than comparing positive vs negative anti-S1 IgG results.

Notably, while IGRA results were highly variable between individuals in our HCW and NISP cohorts, intra-individual results over time remained consistent and predictable, similar to anti-S1 IgG results (eFigure 1A, B).

#### Analysis of solid organ transplant recipients

The immunosuppressive factor screen showed that calcineurin inhibitors, steroids, mycophenolate, and kidney transplant are associated with decreased vaccine response, while liver transplant is associated with high rates of vaccine response. We stratified transplant recipients by their immunosuppressive drug therapy regimens: one drug (12 tacrolimus, 1 everolimus), one drug with steroids (5 tacrolimus, 2 cyclosporine, 1 everolimus, 1 mycophenolate), two drug (18 tacrolimus/mycophenolate, 1 each of sirolimus/mycophenolate, cyclosporine/mycophenolate, and tacrolimus/sirolimus), and two drug with steroids (37 tacrolimus/mycophenolate, 2 tacrolimus/sirolimus, 1 each of cyclosporine/mycophenolate, everolimus/mycophenolate, sirolimus/mycophenolate, tacrolimus/sirolimus, and cyclosporine/azathioprine). Patients on either one-drug or one-drug with steroids regimens were 91% IgG positive (20/22), 86% IgG high (19/22), and 86% IGRA positive (18/21). Comparatively, patients on either two-drug or two-drug with steroids regimens had much lower response rates, at 50% IgG positive (32/64, P<.001), 34% IgG high (22/64, P<.001), and 43% IGRA positive (24/56, P<.001) (eFigure 5A).

To investigate how age affects vaccine response, we examined transplant patients stratified by drug regimen and age (eFigure 5B,C). We chose an age cutoff of 25 based on the age distribution in transplant recipients (eFigure 5B). Patients on antimetabolites who were ≤25 years old were 58% IgG positive (11/19), 47% IgG high (9/19), and 58% IGRA positive 11/19), while those >25 years old were 47% IgG positive (20/43, *P*=.58), 30% IgG high (13/43, *P*=.25), and 34% IGRA positive (12/35, *P*=.15). This shows a potentially slight difference in vaccine response by age.

Next, we examined whether the time between transplant and vaccination affects vaccine response. Patients on antimetabolites who received their transplant more than three years prior to vaccination were 57% IgG positive (27/47), 40% IgG high (19/47), and 50% IGRA positive (20/40), while those who were transplanted within three years were 27% IgG positive (4/15, *P*=.07), 20% IgG high (3/15, *P*=.22), and 21% IGRA positive (3/14, *P*=.11) (eFigure 6A). This shows a potentially slight difference in vaccine response due to the number of years elapsed between transplantation and vaccination.

Results of the immunosuppressive factor screen suggest the transplanted organ is associated with vaccine response. Liver transplant recipients were 100% IgG positive (16/16), 94% IgG high (15/16), and 81% IGRA positive (13/16), while kidney/heart/lung transplant recipients have much lower response rates, at 51% IgG positive (35/69, *P*<.001), 36% IgG high (25/69, *P*<.001), and 47% IGRA positive (28/60, *P*=.02) (eFigure 6B). However, this finding was confounded by the low number (4/16) of liver transplant recipients receiving antimetabolites, and the high number (15/16) receiving their transplant more than three years prior to vaccination (eFigure 6C).

**Analysis of primary immunodeficiency patients**

The immunosuppressive factor screen and multivariable linear regression identified immunoglobulin administration as associated with lower rates of humoral vaccine response. Furthermore, linear regression identified gender as the most predictive factor for vaccine response in patients with primary immunodeficiency. We found that male patients with immunoglobulin deficiency, especially those on immunoglobulin therapy, had low rates of vaccine response (Figure 2C). To rule out differences in different diseases associated with immunoglobulin deficiency (e.g. CVID, hypogammaglobulinemia, hyper-IgM syndrome), we examined only patients diagnosed specifically with CVID. Similarly, compared to NISPs, male CVID patients on immunoglobulin therapy had lower rates of humoral and cellular response, with 25% IgG positive (2/8, *P*<.001), 13% IgG high (1/8, *P*<.001), and 56% IGRA positive (5/9, *P*<.03) (eFigure 7A). Female CVID patients on immunoglobulin therapy had much higher rates of humoral response than their male counterparts, with 100% IgG positive (9/9, *P*=.002) and 78% IgG high (7/9, *P*=.02). Cellular response rates were also higher at 100% IGRA positive (9/9, *P*=.08), albeit this was not statistically significant. Similar age distributions among female and male immunoglobulin deficiency patients suggests that age is not a founding variable (eFigure 7B).

**Analysis of hematologic malignancy patients**

The immunosuppressive factor screen identified active malignancy and B cell lymphoma as predictors of lower rates of humoral and/or cellular response in patients with heme disease. Primary anemia and acute leukemia were predictive of higher rates of humoral and/or cellular response. JAK inhibitors and antimetabolites were identified by multivariable linear regression as factors associated with decreased humoral response. However, as most patients with hematologic malignancies on ISMT received multiple drugs, it is difficult to definitively conclude from our data the effects of specific therapies. Due to small sample sizes, it is difficult to tell if disease activity has strong effect on vaccine response in hematologic malignancy patients. When stratifying patients by both disease activity and ISMT status, the effect of disease activity on anti-S1 IgG and IGRA positivity rates appears to be minimal (eFigure 8A,B).

Multivariable linear regression identified age as the most important predictor of vaccine response in hematologic malignancy patients. However, because of the uneven age distribution of specific malignancies, (eFigure 8C), effects of primary disease versus age were difficult to definitively distinguish.

**Identification of differences in rate of humoral vaccine response decline over time in ISPs**
For anti-S1 IgG, the relationship between rate of change and initial value displays heteroscedasticity, where the variance in the rate of change was higher towards the low end of the initial value (Figure 3B). We hypothesized that this observed heteroscedasticity could be due to two causes: 1) imprecision in the anti-S1 IgG assay, which especially with log transformation, becomes magnified at lower values; 2) certain ISPs with poor humoral response truly have a different antibody titer production dynamics after vaccination, such as having peak titers at greater than 14 days from the time of vaccination. Importantly, ISPs are more likely to be affected by the imprecision of the anti-S1 IgG assay due to the likelihood of having lower anti-S1 IgG values and shorter time between testing compared to HCWs (eFigure 9A).

To identify ISPs with faster or slower rates of decline in humoral response over time than healthy controls (HCWs and NISPs), we determined the 95% CI about the mean rate of change of anti-S1 IgG in HCWs and NISPs combined. ISPs below the lower bound of this CI are labeled as having a faster rate of decline than expected, while those above the upper bound of this CI are labeled as having a slower than expected rate of decline (which includes those with increase vaccine response over time). We then separately analyzed ISPs with a low starting anti-S1 IgG OD ratio (as determined previously) and those with a high initial (or peak) anti-S1 IgG OD ratio, to avoid re-identifying previously identified factors associated with low versus high humoral response.

We performed a screen for immunosuppressive factors that may be associated with a different rate of change of anti-S1 IgG levels over time in ISPs compared to HCWs and NISPs (results not shown). The screen did not identify any immunosuppressive factors that strongly associate with a faster or slower rate of decline, albeit the analysis may be underpowered. Therefore, we attribute most of the heteroscedasticity observed in the data to imprecision in the anti-S1 IgG assay. We conclude that the average change in humoral and cellular vaccine response over time is likely independent of immunosuppression status, both in ISPs with high initial response and low initial response.

**Analysis of booster responses**
We hypothesized that in patients without an anamnestic response to the booster dose, the change in vaccine response before and after the booster dose over the time difference elapse since the most recent vaccine dose will not be significantly different from the change in vaccine response over time after the primary dose. In patients who had a non-anamnestic response, the booster dose simply produced the same response as the primary dose, essentially transporting the patients closer in time to their primary dose, rather than boosting the response (Figure 3D). Therefore, patients with booster response above the response expected at the same timepoint after their primary dose are identified as having an anamnestic response. To account for errors in predicting booster responses, we established confidence intervals around the expected response with errors from equations 2 and 3 (eMethods) when used to predict anti-S1 IgG and IGRA results in individuals with multiple specimens collected after primary vaccination, prior to booster. Reassuringly, the mean prediction error is close to zero for both IgG and IGRA. Also importantly, prediction errors do not correlate with the initial IgG (line of best fit slope = -0.0316, 95% CI -0.0734 to 0.0102, $P$=.14, two-tailed Pearson correlation) and IGRA (line of best fit slope = -0.162, 95% CI -0.347 to 0.0238, $P$=.09, two-tailed Pearson correlation) values (eFigure 9B).

We defined booster responses above the upper bound of the 70% CI of the expected primary dose response as anamnestic. We further defined booster responses above the upper bound of the 95% CI as high anamnestic (eFigure 10A). We suspected that those categorized as having a decreased response (below the lower bound of the 70% CI) after the booster vaccine may be the result of assay imprecision. Therefore, we grouped the decreased response patients with the non-anamnestic response patients.

Of the individuals with paired anti-S1 IgG results available, 66 had a non-anamnestic response to the booster dose, while 18 had an anamnestic response (eFigure 10B). 0/5 HCWs and NISPs and 18/79 ISPs had an anamnestic booster response. Notably, 0/32 ISPs with high anti-S1 IgG levels after primary vaccination had an anamnestic response. This is expected, as these individuals already had "optimal" humoral response after primary vaccination. As noted in the main results section, 18/52 ISPs with low humoral response after primary vaccination had an anamnestic response (Figure 3E).

Of the individuals with paired IGRA results available, 46 had a "non-anamnestic" response, while 2 had an "anamnestic" response (eFigure 10C). 0/6 HCWs and NISPs and 2/42 ISPs had an "anamnestic response."

ISPs not on ISMT had an anamnestic response rate of 19% (3/16), while patients on ISMT had a higher rate of 42% (15/36, P=.13) (eFigure 10D,E). To further identify immunosuppressive factors that predict anamnestic booster response, we performed an immunosuppressive factor screen for anamnestic responders. However, this screen was likely underpowered and did not identify any statistically significant immunosuppressive factors (results not shown).

Three patients on anti-CD20 mAbs had their dose adjusted: two skipped an every-four-month rituximab injection; the third delayed an every-six-month ocrelizumab injection by seven weeks. Only one of these three patients had an anamnestic response, compared to three out of five patients on anti-CD20 mAbs without dose adjustment. Two patients on mycophenolate monotherapy had their dose adjusted: one patient, who was also on tacrolimus and prednisone, had mycophenolate dose decreased from 1250 mg per day to 500 per day for two weeks prior to and after booster vaccination; the other is on mycophenolate monotherapy and had mycophenolate held one week before and two weeks after booster vaccination. Only the latter patient had an anamnestic response. On the other hand, four of seven patients on mycophenolate without dose adjustment had an anamnestic response. Notably, all seven of these patients are also on a calcineurin or mTOR inhibitor with or without steroids. Overall, we find no evidence that temporarily decreasing ISMT dosage or skipping a dose has a significant effect on response to booster vaccination, albeit our sample size is very small.

**eTable 1. Immuno-suppressive/-modulatory therapy (ISMT) counts and categorization for patients in the primary vaccination analysis.**

| Category | n | Specific therapies (n) |
|---|---|---|
| Antimitotics | 13 | Colchicine (1), Docetaxel (2), Vinblastine (2), Vincristine (9), Vinorelbine (1) |
| Tyrosine kinase inhibitors | 6 | Cabozantinib (2), Fostamatinib (1), Nilotinib (1), Sorafenib (2) |
| Other antineoplastics | 15 | Arginine deiminase (1), Bleomycin (2), Bortezomib (1), Carmustine (1), Cisplatin (1), Cyclophosphamide (6), Cytarabine (1), Doxorubicin (8), Etoposide (5), Gemcitabine (2), Palbociclib (1), Panobinostat (1) |
| Calcineurin inhibitors | 100 | Cyclosporine (8), Tacrolimus (93) |
| mTOR inhibitors | 13 | Everolimus (6), Sirolimus (7) |
| Mycophenolate | 79 | Mycophenolate (79) |
| Other antimetabolites | 33 | Azathioprine (9), Hydroxyurea (1), Leflunomide (2), Mercaptopurine (5), Methotrexate (19) |
| Systemic steroids | 105 | Budesonide (2), Dexamethasone (5), Hydrocortisone (1), Methylprednisolone (7), Prednisone (92) |
| Hydroxychloroquine/chloroquine | 17 | Chloroquine (1), Hydroxychloroquine (16) |
| TNFα inhibitors | 10 | Adalimumab (4), Certolizumab (1), Etanercept (2), Infliximab (3) |
| Natalizumab | 3 | Natalizumab (3) |
| Fumarate | 5 | Dimethyl fumarate (3), Diroximel fumarate (2) |
| Other anti-inflammatories | 9 | Apremilast (1), Canakinumab (1), Interferon beta-1a (2), Mesalamine (2), Sulfasalazine (3) |
| Anti-CD20 mAbs | 51 | Ocrelizumab (25), Ofatumumab (1), Rituximab (25) |
| Leukemia/lymphoma targeting | 6 | Acalabritinib (1), Brentuximab vedotin (1), Ibrutinib (2), Pegaspargase (1), Venetoclax (1) |
| S1P receptor modulators | 11 | Fingolimod (8), Ozanimod (1), Siponimod (2) |
| JAK inhibitors | 10 | Baricitinib (1), Ruxolitinib (5), Tofacitinib (4) |
| Tocilizumab | 3 | Tocilizumab (3) |
| Ustekinumab | 4 | Ustekinumab (4) |
| Checkpoint inhibitors | 4 | Avelumab (1), Lenalidomide (2), Pembrolizumab (1) |
| Other immunomodulators | 8 | Abatacept (5), Glatiramer (3) |
| CAR-T* | 5 | CAR-T (5) |
| HSCT | 18 | HSCT within 1 year (18) |
| Immunoglobulins^ | 51 | Immunoglobulins (51) |
| Anti-allergics^ | 5 | Benralizumab (1), Mepolizumab (1), Omalizumab (3) |
| Steroidal hormone targeting^ | 8 | Anastrozole (2), Exemestane (1), Fulvestrant (1), Goserelin (1), Letrozole (2), Mitotane (1), Peptide receptor radionuclide therapy (1) |

*Only CAR-T therapy directed against immune/hematopoietic lineages are included

^Not considered ISMT
Note: for ISMTs dosed greater than four weeks apart, patients were considered to be on the therapy unless they missed a scheduled dose more than four weeks prior to the administration of the first vaccine dose and remained off the therapy until time of testing.

### eTable 2. Immuno-suppressive/-modulatory therapy (ISMT) counts and categorization for patients in the booster vaccination analysis.

| Category | n | Specific therapies (n) |
|---|---|---|
| Antimitotics | 2 | Docetaxel (1), Vincristine (1) |
| Other antineoplastics | 3 | Arginine deiminase (1), Bortezomib (1), Doxorubicin (1), Gemcitabine (1) |
| Calcineurin inhibitors | 32 | Cyclosporine (1), Tacrolimus (31) |
| mTOR inhibitors | 3 | Everolimus (2), Sirolimus (1) |
| Mycophenolate | 25 | Mycophenolate (25) |
| Other antimetabolites | 13 | Azathioprine (4), Leflunomide (2), Mercaptopurine (2), Methotrexate (6) |
| Systemic steroids | 33 | Budesonide (1), Dexamethasone (3), Methylprednisolone (2), Prednisone (28) |
| Hydroxychloroquine/chloroquine | 3 | Hydroxychloroquine (3) |
| TNFα inhibitors | 4 | Adalimumab (2), Etanercept (1), Infliximab (1) |
| Other anti-inflammatories | 2 | Interferon beta-1a (1), Sulfasalazine (1) |
| Anti-CD20 mAbs | 16 | Ocrelizumab (10), Ofatumumab (3), Rituximab (3) |
| Leukemia/lymphoma targeting | 3 | Brentuximab vedotin (1), Ibrutinib (1), Venetoclax (1) |
| S1P receptor modulators | 3 | Fingolimod (3) |
| JAK inhibitors | 2 | Ruxolitinib (1), Tofacitinib (1) |
| Checkpoint inhibitors | 1 | Lenalidomide (1) |
| Other immunomodulators | 2 | Abatacept (2) |
| CAR-T | 3 | CAR-T (3) |
| HSCT | 5 | HSCT within 1 year (5) |
| Immunoglobulins^ | 22 | Immunoglobulins (22) |
| Steroidal hormone targeting^ | 3 | Exemestane (1), Fulvestrant (1), Peptide receptor radionuclide therapy (1) |

*Only CAR-T therapy directed against immune/hematopoietic lineages are included
^Not considered ISMT
Note: for ISMTs dosed greater than four weeks apart, patients were considered to be on the therapy unless they missed a scheduled dose more than four weeks prior to the administration of the first vaccine dose and remained off the therapy until time of testing.

### eTable 3. Disease counts and categorization for patients in the primary vaccination analysis.

| Category/subcategory | n | Specific disease (n) |
|---|---|---|
| Hematologic malignancy | 113 | |
| Plasma cell disease | 7 | Monoclonal gammopathy of unknown significance (3), Monoclonal gammopathy of renal significance (1), Multiple myeloma (2), Waldenstrom macroglobulinemia (1) |
| B cell lymphoma | 57 | Post-transplant lymphoproliferative disorder (B cell morphology) (6), Large B cell lymphoma (13), Follicular lymphoma (9), Hairy cell leukemia (2), Hodgkin lymphoma (6), Lymphoplasmacytic lymphoma (1), MBL/CLL/SLL (9), Mantle cell lymphoma (2), Multicentric Castleman disease (1), Marginal zone lymphoma (6), Primary mediastinal B cell lymphoma (2) |
| T cell Lymphoma | 6 | Anaplastic large cell lymphoma (2), Other T cell lymphoma (2), T cell prolymphocytic leukemia (1), Post-transplant lymphoproliferative disorder (T cell morphology) (1) |
| Acute leukemia | 36 | Acute myeloid leukemia (11), B lymphoblastic leukemia (21), Mixed phenotype acute leukemia (1), T lymphoblastic leukemia (3) |
| MDS & MPN | 10 | Chronic myeloid leukemia (1), Myelodysplastic syndrome (7), Primary myelofibrosis (2) |
| Primary anemia | 9 | Aplastic anemia (4), Beta thalassemia (2), Fanconi anemia (2), Sickle cell disease (1) |
| Solid malignancy | 34 | Adrenocortical carcinoma (2), Bladder carcinoma (1), Breast carcinoma (10), Colorectal carcinoma (1), Ewing sarcoma (2), Germ cell tumor (2), Glioma (1), Hepatoblastoma (1), Hepatocellular carcinoma (1), Leiomyosarcoma (1), Lung carcinoma (1), Meningioma (1), Neuroblastoma (1), Neuroendocrine tumor (1), Osteosarcoma (6), Rhabdomyosarcoma (1), Prostate carcinoma (1), Thyroid carcinoma (1) |
| Solid organ transplant | 88 | Bowel/liver transplant (2), Heart transplant (9), Kidney transplant (52), Liver/kidney transplant (1), Liver transplant (15), Lung transplant (3), Pancreas/kidney transplant (6) |
| Autoimmune disease | 178 | |
| MS/NMO | 58 | Multiple sclerosis (55), Neuromyelitis optica (2), Transverse myelitis (1) |
| Arthritis | 32 | Ankylosing spondylitis (1), Psoriatic arthritis (7), Rheumatoid arthritis (23), Adult Still's disease (1) |
| Lupus/overlap | 34 | Behçet's disease (2), Dermatomyositis (1), Lupus/SLE (18), Mixed connective tissue disease (1), Polymyositis (2), Sjögren's syndrome (7), Systemic sclerosis (3) |
| IBD | 12 | Inflammatory bowel disease (12) |
| Myasthenia gravis | 11 | |
| Good syndrome | 4 | |
| CTLA-4 haploinsufficiency | 3 | |
| Autoimmune hepatitis | 1 | |
| Neuritis | 8 | Autoimmune encephalopathy (2), Autoimmune neuropathy (1), Chronic inflammatory demyelinating polyneuropathy (3), NMDA receptor encephalitis (2) |
| Nephritis | 5 | Goodpasture syndrome (1), IgA nephropathy (4) |

| Category/subcategory | n | Specific disease (n) |
|---|---|---|
| Vasculitis | 7 | ANCA vasculitis (2), Eosinophilic granulomatosis with polyangiitis (1), Giant cell arteritis (1), Granulomatosis with polyangiitis (2), Microscopic polyangiitis (1) |
| Dermatitis | 4 | Atopic dermatitis (2), Chronic dermatitis (1), Lichen sclerosus (1) |
| Autoimmune thyroiditis | 3 | |
| Other autoimmune disease | 7 | Antisynthetase syndrome (2), IgG4-related disease (1), Myalgic encephalomyelitis (1), Polyglandular autoimmune syndrome type 1 (1), Systemic inflammatory response syndrome (1), STAT3 gain-of-function mutation (1) |
| 1° Immunodeficiency | 49 | |
| Immunoglobulin deficiency | 41 | Common variable immunodeficiency (30), Hyper-IgM syndrome (2), Hypogammaglobulinemia (8), IgA deficiency (1) |
| Other immunodeficiency | 9 | 22q11.2 deletion syndrome (2), Anti-interferon-γ immunodeficiency syndrome (1), CD4 lymphopenia (3), GATA2 mutation (1), McKusick cartilage-hair hypoplasia associated with B & T lymphopenia (1), Severe combined immunodeficiency (1) |

Note: counts include ISPs with diseases belonging to multiple categories

### eTable 4. Disease counts and categorization for patients in the booster vaccination analysis.

| Category/subcategory | n | Specific disease (n) |
|---|---|---|
| Hematologic malignancy | 39 | |
| Plasma cell disease | 4 | Monoclonal gammopathy of unknown significance (2), Multiple myeloma (1), Waldenstrom macroglobulinemia (1) |
| B cell lymphoma | 18 | Post-transplant lymphoproliferative disorder (B cell morphology) (1), Large B cell lymphoma (5), Follicular lymphoma (2), Hodgkin lymphoma (1), MBL/CLL/SLL (6), Mantle cell lymphoma (1), Marginal zone lymphoma (2) |
| T cell Lymphoma | 4 | Anaplastic large cell lymphoma (1), Other T cell lymphoma (2), T cell prolymphocytic leukemia (1) |
| Acute leukemia | 9 | Acute myeloid leukemia (5), B lymphoblastic leukemia (4) |
| MDS & MPN | 5 | Myelodysplastic syndrome (5) |
| Solid malignancy | 4 | Breast carcinoma (3), Leiomyosarcoma (1), Neuroendocrine tumor (1) |
| Solid organ transplant | 30 | Bowel/liver transplant (1), Heart transplant (1), Kidney transplant (21), Liver/kidney transplant (1), Liver transplant (3), Lung transplant (1), Pancreas/kidney transplant (2) |
| Autoimmune disease | 48 | |
| MS/NMO | 18 | Multiple sclerosis (18) |
| Arthritis | 7 | Ankylosing spondylitis (1), Rheumatoid arthritis (6) |
| Lupus/overlap | 10 | Lupus/SLE (6), Polymyositis (2), Sjögren's syndrome (1), Systemic sclerosis (1) |
| IBD | 3 | |
| Myasthenia gravis | 3 | |
| Good syndrome | 1 | |
| Autoimmune hepatitis | 1 | |
| Neuritis | 1 | Chronic inflammatory demyelinating polyneuropathy (1) |
| Nephritis | 1 | IgA nephropathy (1) |
| Vasculitis | 2 | ANCA vasculitis (2) |
| Dermatitis | 1 | Lichen sclerosus (1) |
| Other autoimmune disease | 4 | IgG4-related disease (1), Myalgic encephalomyelitis (1), Sarcoidosis (2) |
| 1° Immunodeficiency | 18 | |
| Immunoglobulin deficiency | 16 | Common variable immunodeficiency (14), Hypogammaglobulinemia (1), IgA deficiency (1) |
| Other immunodeficiency | 2 | CD4 lymphopenia (2) |

Note: counts include ISPs with diseases belonging to multiple categories

**eTable 5. Availability of IFN-γ release assay (IGRA) and Anti-S1 IgG (IgG) assay results in cohorts.**

| | | HCW | NISP | ISP |
|---|---|---|---|---|
| After primary vaccination | IGRA and IgG | 18 | 25 | 381 |
| | IGRA only | 0 | 1 | 8 |
| | IgG only | 0 | 2 | 38 |
| Multiple assay results after primary vaccination | IGRA and IgG | 17 | 1 | 16 |
| | IGRA only | 0 | 0 | 0 |
| | IgG only | 0 | 3 | 54 |
| After booster | IGRA and IgG | 6 | 5 | 117 |
| | IGRA only | 0 | 1 | 7 |
| | IgG only | 0 | 0 | 1 |
| Before and after booster vaccination | IGRA and IgG | 6 | 0 | 53 |
| | IGRA only | 0 | 0 | 0 |
| | IgG only | 0 | 1 | 31 |

### eTable 6. Primary vaccine count in cohorts.

| | HCW | NISP | ISP |
|---|---|---|---|
| BNT162b2 | 18 | 14 | 269 |
| mRNA-1273 | 0 | 13 | 142 |
| Mixed* | 0 | 1 | 4 |
| Ad26.COV2.S | 0 | 0 | 12 |

*Mixed: a mixture of one dose of BNT162b2 and one dose of mRNA-1273 (in either order)

**eTable 7. Assay positivity rate in cohorts with both IFN-γ release assay (IGRA) and anti-S1 IgG (IgG) results available after primary vaccination.**

| | | | | Disease Categories | | | | | | | |
|---|---|---|---|---|---|---|---|---|---|---|---|
| | **HCW (n=18)** | **NISP (n=25)** | **ISP (n=381)** | **Active Heme Malignancy (n=25)** | **Inactive Heme Malignancy (n=55)** | **Primary anemia (n=9)** | **Solid Malignancy (n=22)** | **Solid Organ Transplant (n=59)** | **Autoimmune Disease (n=132)** | **1° Immuno-deficiency (n=39)** | **Multiple categories (n=40)** |
| Both IGRA and IgG positive % (n) | 100 (18) | 92 (23) | 51 (196) | 32 (8) | 64 (35) | 89 (8) | 77 (17) | 51 (30) | 46 (61) | 62 (24) | 32 (13) |
| Both IGRA and IgG negative % (n) | 0 (0) | 0 (0) | 18 (67) | 24 (6) | 13 (7) | 0 (0) | 9 (2) | 29 (17) | 17 (22) | 5 (2) | 28 (11) |
| IGRA positive and IgG negative % (n) | 0 (0) | 0 (0) | 20 (75) | 24 (6) | 16 (9) | 11 (1) | 5 (1) | 7 (4) | 31 (41) | 15 (6) | 18 (7) |
| IgG positve and IGRA negative % (n) | 0 (0) | 8 (2) | 11 (43) | 20 (5) | 7 (4) | 0 (0) | 9 (2) | 14 (8) | 6 (8) | 18 (7) | 22 (9) |
| IGRA positive and IgG high % (n) | 100 (18) | 88 (22) | 42 (160) | 16 (4) | 53 (29) | 89 (8) | 77 (17) | 42 (25) | 36 (47) | 49 (19) | 28 (11) |
| IGRA negative and IgG low % (n) | 0 (0) | 4 (1) | 24 (92) | 36 (9) | 16 (9) | 0 (0) | 18 (4) | 36 (21) | 20 (26) | 21 (8) | 38 (15) |
| IGRA positive and IgG low % (n) | 0 (0) | 4 (1) | 29 (111) | 40 (10) | 27 (15) | 11 (1) | 5 (1) | 15 (9) | 42 (55) | 28 (11) | 22 (9) |
| IgG high and IGRA negative % (n) | 0 (0) | 4 (1) | 5 (18) | 8 (2) | 4 (2) | 0 (0) | 0 (0) | 7 (4) | 3 (4) | 3 (1) | 12 (5) |

### eTable 8. Immunosuppressive factors associated with low anti-S1 IgG (*P*<.05) after primary vaccination.

| Immunosuppressive factor[&] | Odds ratio of low IgG[#] | Affected % high IgG (n)^ | Not affected % high IgG (n)[§] | Fisher exact *P* value |
|---|---|---|---|---|
| Anti-CD20 mAbs | 8.31 | 12.5 (6/48) | 54.3 (216/398) | <.001* |
| Ocrelizumab | 11.98 | 8.3 (2/24) | 52.1 (220/422) | <.001* |
| Autoimmune disease | 2.16 | 38.2 (66/173) | 57.1 (156/273) | <.001* |
| S1P receptor modulators | indeterminate | 0.0 (0/11) | 51.0 (222/435) | <.001* |
| Systemic steroids | 2.09 | 35.9 (37/103) | 53.9 (185/343) | .002* |
| Mycophenolate | 2.28 | 33.3 (26/78) | 53.3 (196/368) | .002* |
| Active heme malignancy | 3.51 | 23.5 (8/34) | 51.9 (214/412) | .002* |
| Rituximab | 5.05 | 17.4 (4/23) | 51.5 (218/423) | .002* |
| Multiple sclerosis | 2.63 | 29.6 (16/54) | 52.6 (206/392) | .002* |
| B cell lymphoma | 2.46 | 30.9 (17/55) | 52.4 (205/391) | .004* |
| Fingolimod | indeterminate | 0.0 (0/8) | 50.7 (222/438) | .007* |
| Immunoglobulins | 2.30 | 32.0 (16/50) | 52.0 (206/396) | .01* |
| MS/NMO | 2.18 | 33.3 (19/57) | 52.2 (203/389) | .01* |
| Prednisone | 1.84 | 37.8 (34/90) | 52.8 (188/356) | .01* |
| Kidney transplant | 2.03 | 34.6 (18/52) | 51.8 (204/394) | .03 |
| Marginal zone lymphoma | indeterminate | 0.0 (0/6) | 50.5 (222/440) | .03 |
| JAK inhibitors | 8.19 | 11.1 (1/9) | 50.6 (221/437) | .04 |
| B cell lineage affected | 1.56 | 42.0 (55/131) | 53.0 (167/315) | .04 |

[&] Specific disease, disease subcategory, disease category, specific therapy, therapy category, or immune lineages affected by disease (myeloid, B cell, and T cell). Note: only immunosuppressive factors with n ≥ 3 affected patients are included in the analysis
[#] Odds ratio that patients affected by the immunosuppressive factor have low anti-S1 IgG. OR>1 indicates that the factor is associated with increased likelihood of having low IgG
^ % of patients affected by the immunosuppressive factor that have high anti-S1 IgG
[§] % of patients not affected by the immunosuppressive factor that have high anti-S1 IgG
* Statistically significant *P* value after correction for multiple comparisons

### eTable 9. Immunosuppressive factors associated with high anti-S1 IgG (top 10 by *P* value) after primary vaccination.

| Immunosuppressive factor[&] | Odds ratio of low IgG[#] | Affected % high IgG (n)^ | Not affected % high IgG (n)[§] | Fisher exact *P* value |
|---|---|---|---|---|
| Not on any therapy | 0.27 | 72.5 (87/120) | 41.4 (135/326) | <.001* |
| Not immunosuppressed | 0.07 | 92.6 (25/27) | 47.0 (197/419) | <.001* |
| Liver transplant | 0.07 | 93.3 (14/15) | 48.3 (208/431) | <.001* |
| Solid malignancy | 0.28 | 76.5 (26/34) | 47.6 (196/412) | .001* |
| Primary anemia | 0.12 | 88.9 (8/9) | 49.0 (214/437) | .02 |
| Other anti-inflammatories | 0.12 | 88.9 (8/9) | 49.0 (214/437) | .02 |
| Fumarate | 0 | 100.0 (5/5) | 49.2 (217/441) | .03 |
| B lymphoblastic leukemia | 0.38 | 71.4 (15/21) | 48.7 (207/425) | .05 |
| Neuritis | 0.16 | 85.7 (6/7) | 49.2 (216/439) | .07 |
| Acute leukemia | 0.53 | 63.9 (23/36) | 48.5 (199/410) | .08 |

[&] Specific disease, disease subcategory, disease category, specific therapy, therapy category, or immune lineages affected by disease (myeloid, B cell, and T cell). Note: only immunosuppressive factors with n ≥ 3 affected patients are included in the analysis
[#] Odds ratio that patients affected by the immunosuppressive factor have low anti-S1 IgG. OR>1 indicates that the factor is associated with increased likelihood of having low IgG
^ % of patients affected by the immunosuppressive factor that have high anti-S1 IgG
[§] % of patients not affected by the immunosuppressive factor that have high anti-S1 IgG
* Statistically significant *P* value after correction for multiple comparisons

### eTable 10. Immunosuppressive factors associated with negative IFN-γ response (*P*<.05) after primary vaccination.

| Immunosuppressive factor[&] | Odds ratio of neg IGRA[#] | Affected % pos IGRA (n)^ | Not affected % pos IGRA (n)[§] | Fisher exact *P* value |
|---|---|---|---|---|
| Calcineurin inhibitors | 3.81 | 49.4 (44/89) | 78.8 (257/326) | <.001* |
| Systemic steroids | 3.44 | 51.6 (48/93) | 78.6 (253/322) | <.001* |
| Tacrolimus | 3.46 | 50.6 (42/83) | 78.0 (259/332) | <.001* |
| Prednisone | 3.43 | 50.6 (41/81) | 77.8 (260/334) | <.001* |
| S1P receptor modulators | 28.85 | 9.1 (1/11) | 74.3 (300/404) | <.001* |
| Solid organ transplant | 2.96 | 53.2 (42/79) | 77.1 (259/336) | <.001* |
| Mycophenolate | 2.58 | 55.1 (38/69) | 76.0 (263/346) | <.001* |
| Fingolimod | 19.63 | 12.5 (1/8) | 73.7 (300/407) | <.001* |
| Kidney transplant | 3.03 | 50.0 (22/44) | 75.2 (279/371) | .001* |
| T cell lineage affected | 3.39 | 45.8 (11/24) | 74.2 (290/391) | .004* |
| Mercaptopurine | indeterminate | 0.0 (0/4) | 73.2 (301/411) | .006* |
| Antimitotics | 4.47 | 38.5 (5/13) | 73.6 (296/402) | .009* |
| Cyclosporine | 6.86 | 28.6 (2/7) | 73.3 (299/408) | .02 |
| Lung transplant | indeterminate | 0.0 (0/3) | 73.1 (301/412) | .02 |
| Dexamethasone | 10.91 | 20.0 (1/5) | 73.2 (300/410) | .02 |
| Other antimetabolites | 2.45 | 53.6 (15/28) | 73.9 (286/387) | .03 |
| Active heme malignancy | 2.20 | 56.2 (18/32) | 73.9 (283/383) | .04 |

[&] Specific disease, disease subcategory, disease category, specific therapy, therapy category, or immune lineages affected by disease (myeloid, B cell, and T cell). Note: only immunosuppressive factors with n ≥ 3 affected patients are included in the analysis
[#] Odds ratio that patients affected by the immunosuppressive factor have negative IFN-γ response. OR>1 indicates that the factor is associated with increased likelihood of having negative IFN-γ response
^ % of patients affected by the immunosuppressive factor that have positive IFN-γ response
[§] % of patients not affected by the immunosuppressive factor that have positive IFN-γ response
* Statistically significant *P* value after correction for multiple comparisons

**eTable 11. Immunosuppressive factors associated with positive IFN-γ response (top 10 by *P* value) after primary vaccination.**

| Immunosuppressive factor[&] | Odds ratio of neg IGRA[#] | Affected % pos IGRA (n)^ | Not affected % pos IGRA (n)[§] | Fisher exact *P* value |
|---|---|---|---|---|
| Not on any therapy | 0.25 | 88.6 (101/114) | 66.4 (200/301) | <.001* |
| Ocrelizumab | 0.11 | 95.8 (23/24) | 71.1 (278/391) | .008* |
| Not immunosuppressed | 0.21 | 92.3 (24/26) | 71.2 (277/389) | .02 |
| Anti-CD20 mAbs | 0.48 | 83.7 (41/49) | 71.0 (260/366) | .09 |
| TNFα inhibitors | 0 | 100.0 (8/8) | 72.0 (293/407) | .11 |
| Primary anemia | 0 | 100.0 (9/9) | 71.9 (292/406) | .12 |
| Other autoimmune disease | 0 | 100.0 (6/6) | 72.1 (295/409) | .19 |
| Multiple sclerosis | 0.66 | 79.2 (42/53) | 71.5 (259/362) | .32 |
| Fumarate | 0 | 100.0 (5/5) | 72.2 (296/410) | .33 |
| MS/NMO | 0.69 | 78.6 (44/56) | 71.6 (257/359) | .34 |

[&] Specific disease, disease subcategory, disease category, specific therapy, therapy category, or immune lineages affected by disease (myeloid, B cell, and T cell). Note: only immunosuppressive factors with n ≥ 3 affected patients are included in the analysis
[#] Odds ratio that patients affected by the immunosuppressive factor have negative IFN-γ response. OR>1 indicates that the factor is associated with increased likelihood of having negative IFN-γ response
^ % of patients affected by the immunosuppressive factor that have positive IFN-γ response
[§] % of patients not affected by the immunosuppressive factor that have positive IFN-γ response
* Statistically significant *P* value after correction for multiple comparisons

**eTable 12. Multivariable linear regression with dependent variable anti-S1 IgG OD ratio after primary vaccination.**

| **Cohort** | **NISP+ISP** | **Heme Disease (heme malignancy and primary anemia)** | **Solid Malignancy** | **Solid Organ Transplant** | **Autoimmune Disease** | **1º Immuno-deficiency** |
|---|---|---|---|---|---|---|
| **Independent (predictor) variables** | Base predictors* | Base predictors* + HSCT within 5 years + HSCT within 10 years | Base predictors* | Base predictors* + years since transplant | Base predictors* | Base predictors* |
| **n** | 446 | 101[&] | 24[&] | 88[&] | 144[&] | 41[&] |
| **Adjusted $R^2$** | 0.40 | 0.45 | -0.09 | 0.36 | 0.42 | 0.28 |
| **F statistic** | 5.34 (*P*<.001) | 4.00 (*P*<.001) | 0.79 (*P*=.63) | 3.61 (*P*<.001) | 4.10 (*P*<.001) | 3.17 (*P*=.01) |
| **Significant predictors^ (coefficient)** | Mycophenolate (-3.27, *P*<.001) | Age (-0.07, *P*=.003) | | Mycophenolate (-2.93, *P*=.003) | Anti-CD20 mAbs (-3.75, *P*<.001) | Gender (-2.34, *P*=.02) |
| | Anti-CD20 mAbs (-3.36, *P*<.001) | JAK inhibitors (-4.34, *P*=.01) | | B cell lymphoma (B cell morphology PTLD, -2.40, *P*=.01) | Mycophenolate (-4.35, *P*<.001) | Immunoglobulin therapy (-2.12, *P*=.03) |
| | S1P receptor modulators (-3.91, *P*<.001) | Other antimetabolites (-3.17, *P*=.04) | | | S1P receptor modulators (-4.46, *P*<.001) | |
| | mTOR inhibitors (-3.28, *P*<.001) | | | | On any ISMT (2.47, *P*=.008) | |
| | JAK inhibitors (-3.57, *P*<.001) | | | | Immunoglobulin therapy (2.46, *P*=.01) | |
| | Days since vaccine (-0.01, *P*<.001) | | | | Arthritis (-2.03, *P*=.04) | |
| | Neuritis (4.05, *P*=.004) | | | | Other autoimmune disease (-2.83, *P*=.05) | |
| | Other antimetabolites (-1.90, *P*=.004) | | | | Systemic steroids (-1.21, *P*=.05) | |
| | Age (-0.02, *P*=.005) | | | | | |
| | Gender (-0.68, *P*=.02) | | | | | |
| | Calcineurin inhibitors (-1.71, *P*=.02) | | | | | |

| Cohort | NISP+ISP | Heme Disease (heme malignancy and primary anemia) | Solid Malignancy | Solid Organ Transplant | Autoimmune Disease | 1º Immuno-deficiency |
|---|---|---|---|---|---|---|
| **Significant predictors^ (coefficient)** | Plasma cell disease (-3.76, *P*=.03) | | | | | |
| | Hydroxychloroquine/ chloroquine (-1.85, *P*=.03) | | | | | |
| | Ustekinumab (-4.12, *P*=.03) | | | | | |
| | On any ISMT (1.40, *P*=.03) | | | | | |
| | Leukemia/ lymphoma targeting therapy (-2.74, *P*=.03) | | | | | |

* Base predictors: age, gender, days since vaccine administration, immune lineage affected by primary disease (myeloid, B, or T), disease categories, disease subcategories, therapy categories, and on any ISMT
^ Significant predictors: predictors with statistically significant coefficient (*P*<.05), ranked by *P* value; only reported if model F statistic is significant (*P*<.05)
[&] Includes all ISPs with a disease belonging to the category, including those with diseases belonging to multiple categories

### eTable 13. Multivariable linear regression with dependent variable IFN-γ response (IU/mL) after primary vaccination.

| Cohort | NISP+ISP | Heme Disease (heme malignancy and primary anemia) | Solid Malignancy | Solid Organ Transplant | Autoimmune Disease | 1º Immunodeficiency |
|---|---|---|---|---|---|---|
| **Independent (predictor) variables** | Base predictors* | Base predictors* + HSCT within 5 years + HSCT within 10 years | Base predictors* | Base predictors* + years since transplant | Base predictors* | Base predictors* |
| **n** | 415 | 91[&] | 22[&] | 79[&] | 137[&] | 40[&] |
| **Adjusted $R^2$** | 0.15 | -0.02 | -0.08 | 0.14 | 0.05 | 0.19 |
| **F statistic** | 2.03 (*P*<.001) | 0.95 (*P*=.54) | 0.84 (*P*=.60) | 1.69 (*P*=.07) | 1.22 (*P*=.22) | 2.30 (*P*=.05) |
| **Significant predictors^ (coefficient)** | Anti-CD20 mAbs (2.45, *P*=.002) | | | | | |
| | CAR-T (4.49, *P*=.01) | | | | | |
| | TNFα inhibitors (3.00, *P*=.03) | | | | | |
| | Age (-0.02, *P*=.03) | | | | | |
| | On any ISMT (-1.77, *P*=.04) | | | | | |

* Base predictors: age, gender, days since vaccine administration, immune lineage affected by primary disease (myeloid, B, or T), disease categories, disease subcategories, therapy categories, and on any ISMT
^ Significant predictors: predictors with statistically significant coefficient (*P*<.05), ranked by *P* value; only reported if model F statistic is significant (*P*<.05)
[&] Includes all ISPs with a disease belonging to the category, including those with diseases belonging to multiple categories

**eTable 14. Booster vaccine count in cohorts.**

| | HCW | NISP | ISP |
|---|---|---|---|
| BNT162b2 | 6 | 4 | 75 |
| mRNA-1273 | 0 | 2 | 47 |
| Ad26.COV2.S | 0 | 0 | 3 |
| Booster matches primary vaccine | 6 | 5 | 117 |
| Booster does not match primary vaccine | 0 | 1 | 8 |
| Median time between primary and booster doses (IQR) | 259.5 (252-270) | 185 (157.8-225.8) | 144 (123.0-172.3) |

**eTable 15. Assay positivity rate in cohorts with both IFN-γ release assay (IGRA) and anti-S1 IgG (IgG) results available after booster.**

| | | | | Disease Categories | | | | | | |
|---|---|---|---|---|---|---|---|---|---|---|
| | **HCW (n=6)** | **NISP (n=5)** | **ISP (n=117)** | **Active Heme Malignancy (n=10)** | **Inactive Heme Malignancy (n=20)** | **Solid Malignancy (n=2)** | **Solid Organ Transplant (n=23)** | **Autoimmune Disease (n=34)** | **1° Immuno-deficiency (n=17)** | **Multiple categories (n=11)** |
| Both IGRA and IgG positive % (n) | 100 (6) | 80 (4) | 55 (64) | 50 (5) | 45 (9) | 100 (2) | 48 (11) | 62 (21) | 71 (12) | 36 (4) |
| Both IGRA and IgG negative % (n) | 0 (0) | 0 (0) | 14 (16) | 20 (2) | 15 (3) | 0 (0) | 22 (5) | 9 (3) | 0 (0) | 27 (3) |
| IGRA positive and IgG negative % (n) | 0 (0) | 0 (0) | 18 (21) | 30 (3) | 30 (6) | 0 (0) | 9 (2) | 21 (7) | 12 (2) | 9 (1) |
| IgG positive and IGRA negative % (n) | 0 (0) | 20 (1) | 14 (16) | 0 (0) | 10 (2) | 0 (0) | 22 (5) | 9 (3) | 18 (3) | 27 (3) |

**eFigure 1. Determination of expected decline in vaccine response over time in HCWs.**

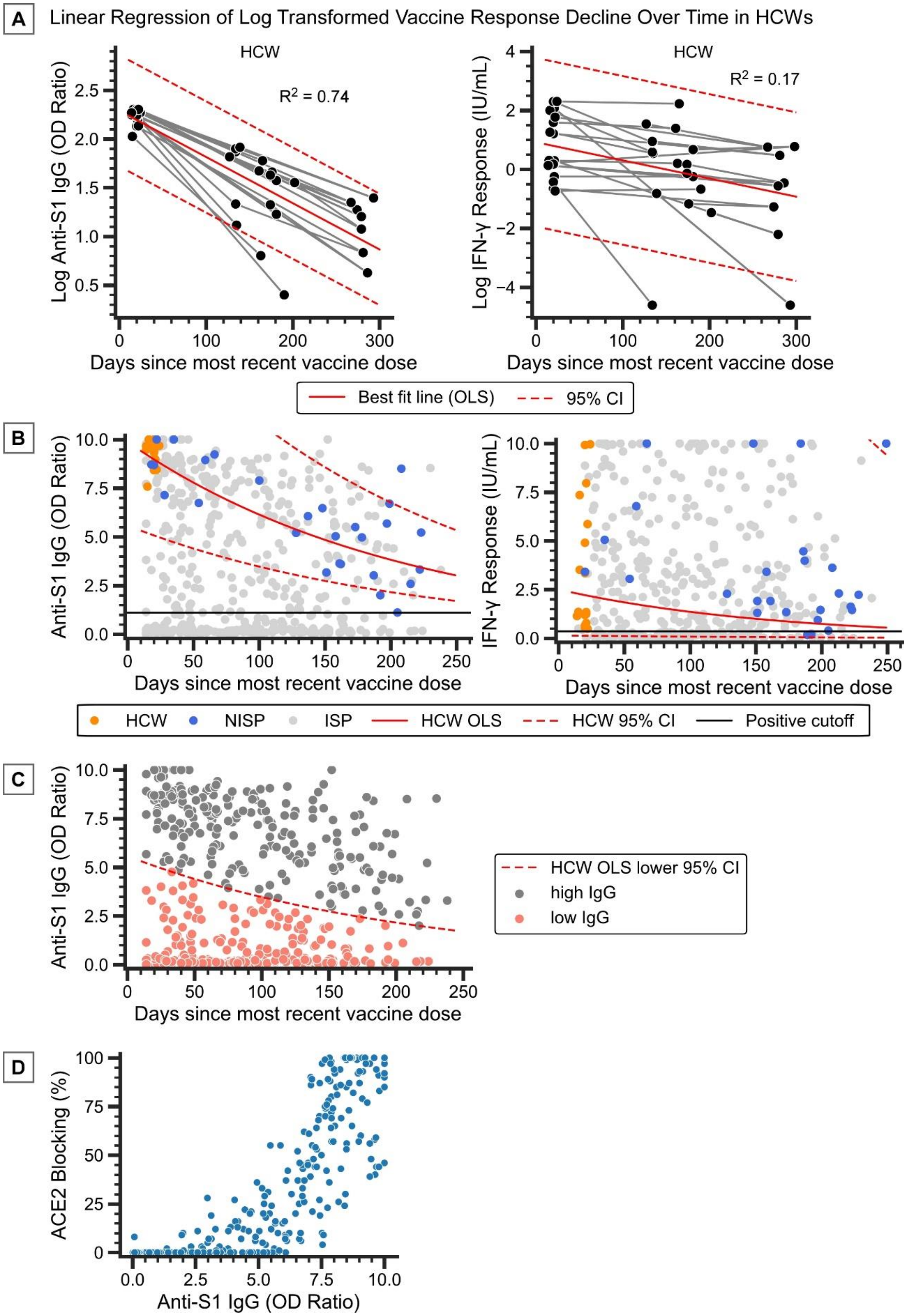

A) Linear regression models fitted to the change over time of natural log transformed anti-S1 IgG and IFN-γ response in the HCW cohort, with 95% confidence intervals (CI) calculated based on the standard deviation of the residuals. Models were calculated using ordinary least squares (OLS), with $R^2$ as indicated. Gray lines connect repeat assay results from the same individual. Note, plotted are all anti-S1 IgG and IGRA results from 18 HCWs after primary vaccination (prior to booster vaccination). One of the HCWs has one of each result available, while seven have three anti-S1 IgG results available collected at different times after vaccination, and eight have three IGRA results available. For the linear regression, only the 17 HCWs with at least two anti-S1 IgG and IGRA results were included, and only two datapoints were included for each HCW (the two furthest apart in time of collection).

B) Linear regression models from panel A plotted with non-log transformed anti-S1 IgG and IFN-γ response values. Shown also are NISP and ISP cohorts.

C) Definition of higher and lower than expected anti-S1 IgG (high IgG and low IgG) in NISP and ISP cohorts based on the lower bound of the 95% CI calculated using equation 1 (eMethods).

D) Correlation of ACE2 blocking % with anti-S1 IgG OD ratio in HCWs, NISPs, and ISPs.

**eFigure 2. Distribution of anti-S1 IgG, ACE2 blocking, and IFN-γ response values in the HCW, NISP, and ISP cohorts categorized by disease and therapy status.**

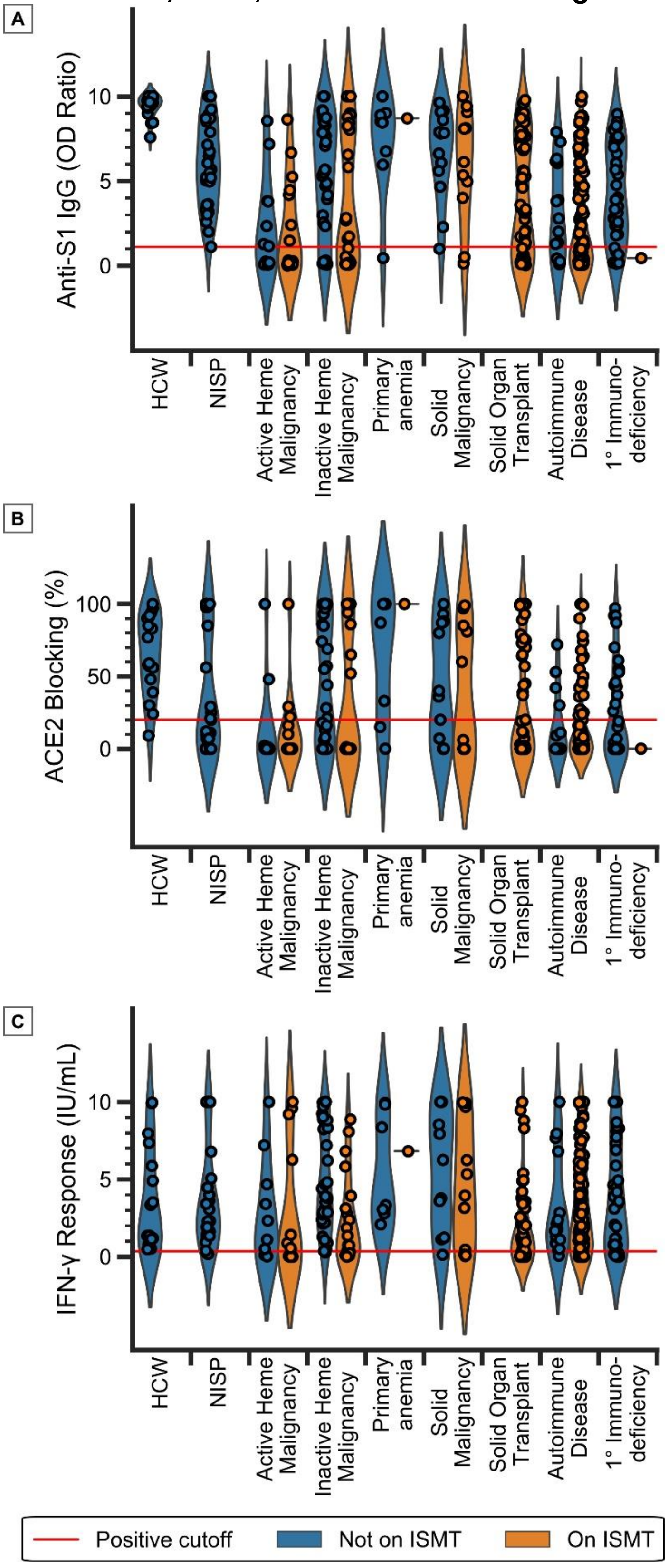

Only patients with diseases belonging to a single category are plotted. Red lines represent the positive cutoff for each respective assay.

**eFigure 3. Immunosuppressive factors associated with low and high anti-S1 IgG in NISP and ISP cohorts.**

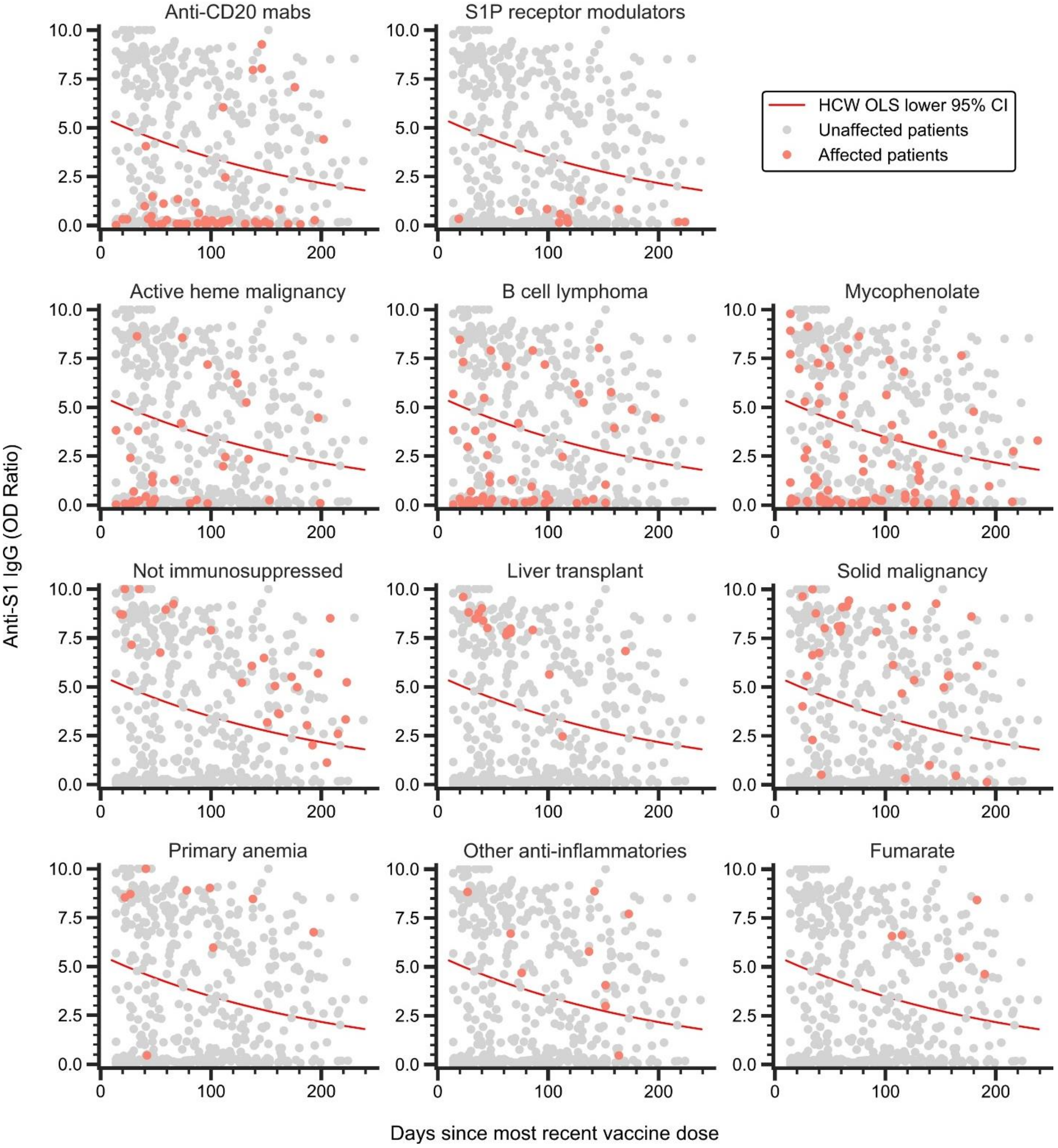

Results of the immunosuppressive factor screen (eTables 8,9). Red line indicates the lower bound of the 95% confidence interval (CI) calculated using equation 1 (eMethods), which separates high anti-S1 IgG from low anti-S1 IgG. Affected/unaffected patients refer to patients affected/unaffected by the immunosuppressive factor indicated.

**eFigure 4. Immunosuppressive factors associated with negative and positive IFN-γ response in NISP and ISP cohorts.**

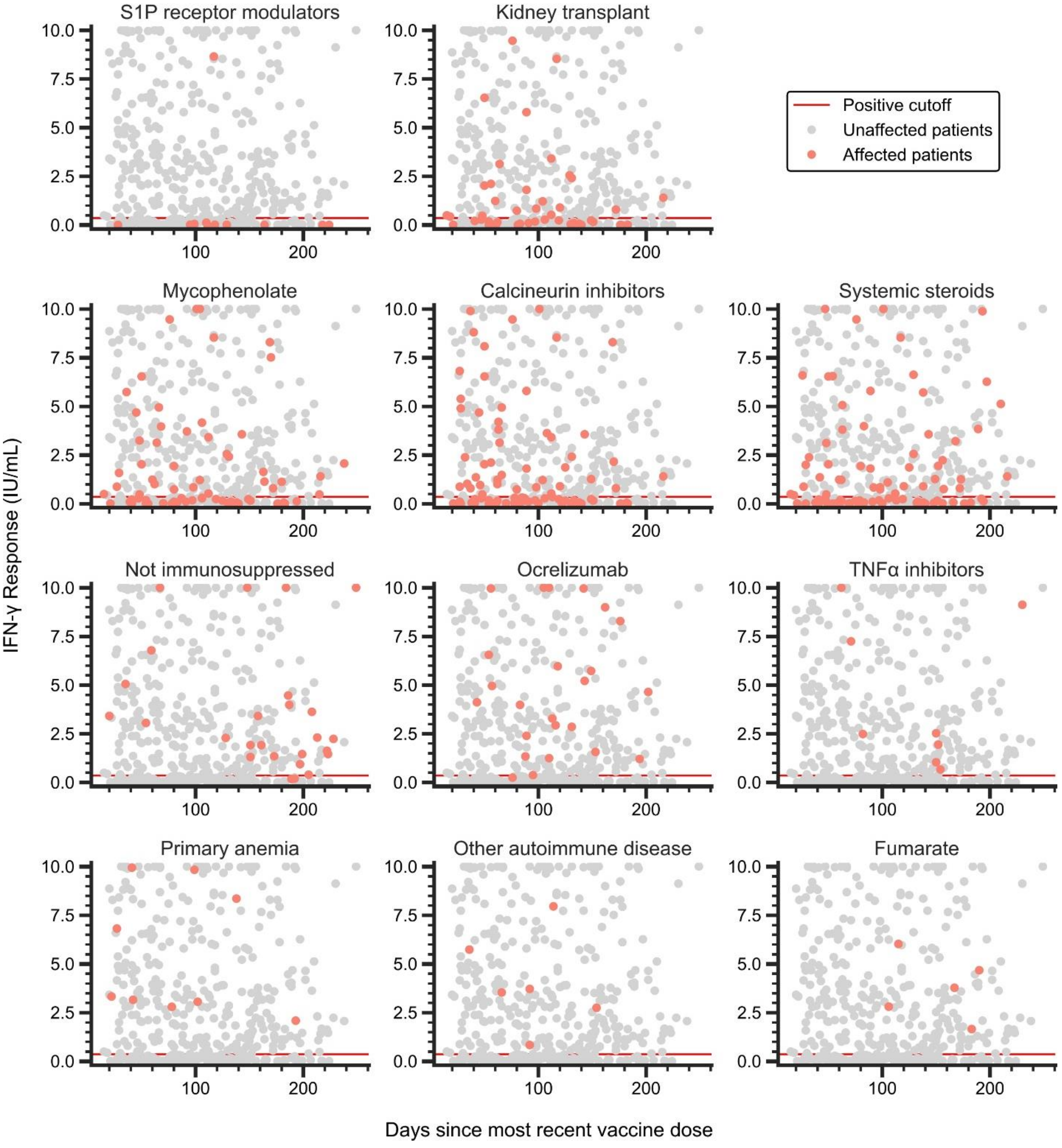

Results of the immunosuppressive factor screen (eTables 10,11). Red line indicates the positive cutoff for IGRA. Affected/unaffected patients refer to patients affected/unaffected by the immunosuppressive factor indicated.

**eFigure 5. Age is a potential weak predictor of humoral and cellular vaccine response in solid organ transplant recipients.**

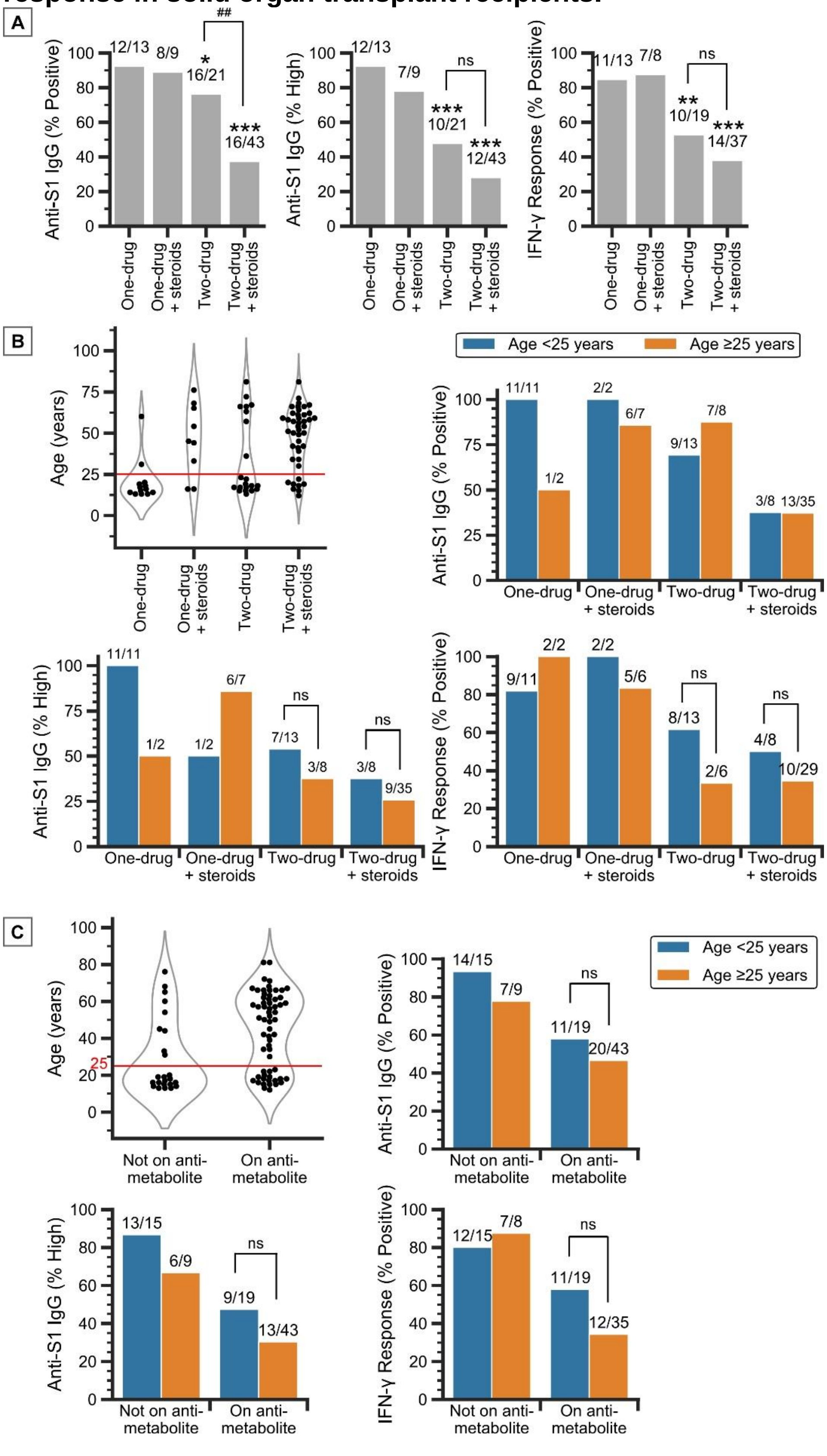

A) Vaccine response rate in transplant recipients on one-drug, one-drug with steroids, two-drug, and two-drug with steroids regimens.

B) Age distribution of transplant recipients on one-drug, one-drug with steroids, two-drug, and two-drug with steroids regimens shows a bimodal distribution above and below age 25. Vaccine response positivity rates of transplant recipients stratified by drug regimen and age (<25 and ≥25 years).

C) Age distribution of transplant recipients not on and on antimetabolites shows a bimodal distribution above and below age 25. Vaccine response positivity rates of transplant recipients stratified by antimetabolite use and age (<25 and ≥25 years).

Only transplant recipients on a calcineurin inhibitor, mTOR inhibitor, mycophenolate, or other antimetabolites are plotted. ns, not significant; $^{\#}P$<.05, $^{\#\#}P$<.01, $^{\#\#\#}P$<.001 for pairwise comparisons as indicated; **P*<.05, ***P*<.01, ****P*<.001 compared to NISP, Fisher exact test.

**eFigure 6. Years between solid organ transplant and vaccination is a potential weak predictor of humoral and cellular vaccine response in transplant recipients.**

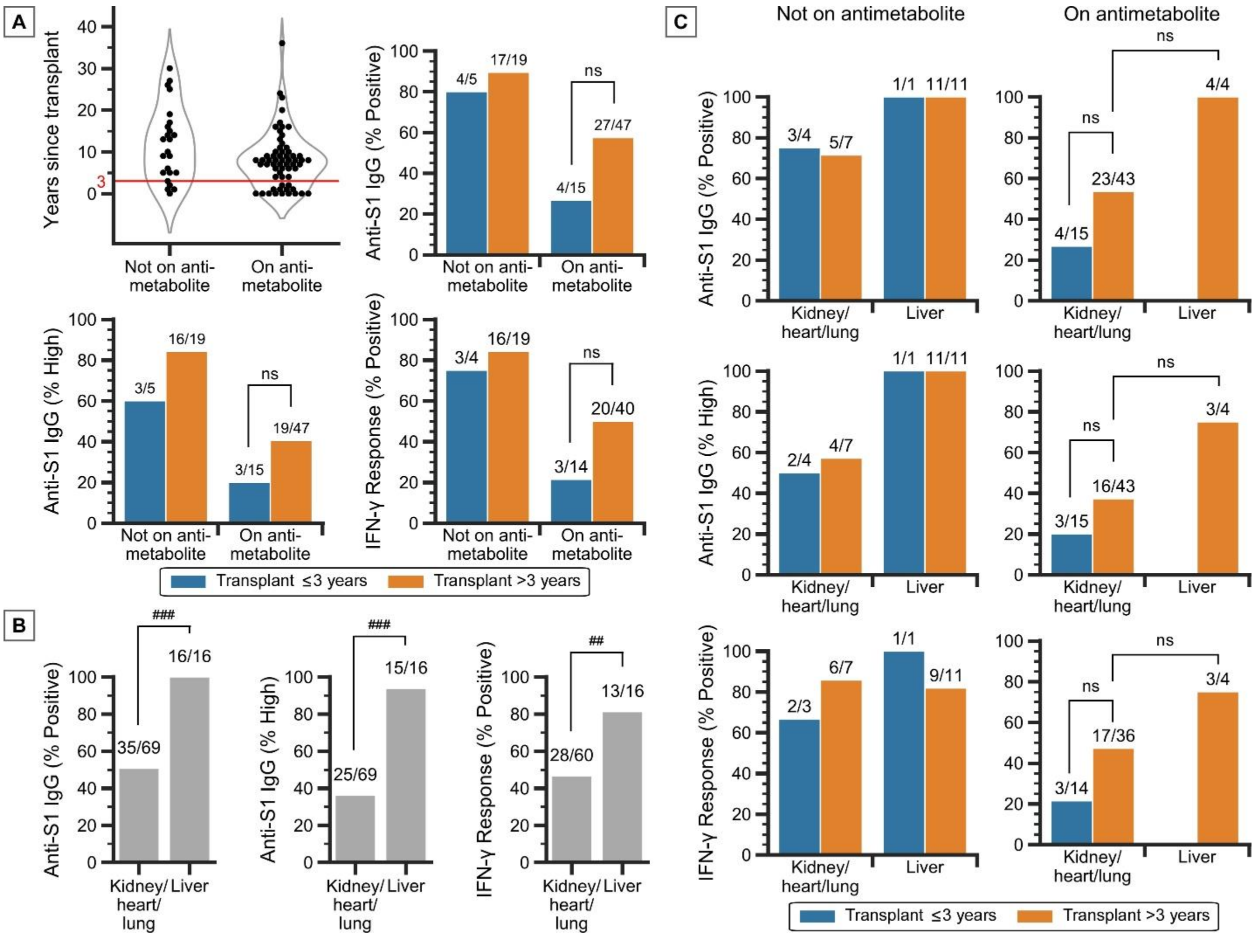

A) Distribution of years between transplant and vaccination in transplant recipients not on and on antimetabolites shows a bimodal pattern above and below three years. Vaccine response positivity rates of transplant recipients stratified by antimetabolite use and years since transplant (<3 and ≥3).

B) Vaccine response positivity rates of transplant recipients stratified by organ transplanted (kidney/heart/lung and liver).

C) Vaccine response positivity rates of transplant recipients stratified by organ transplanted and years since transplant (<3 and ≥3). Those not on antimetabolites are presented on the left column; those on antimetabolites are presented on the right column.

Only transplant recipients on a calcineurin inhibitor, mTOR inhibitor, mycophenolate, or other antimetabolites are plotted. ns, not significant; $^{\#}P$<.05, $^{\#\#}P$<.01, $^{\#\#\#}P$<.001, Fisher exact test.

### eFigure 7. Male gender is associated with decreased vaccine response in CVID patients on immunoglobulin therapy.

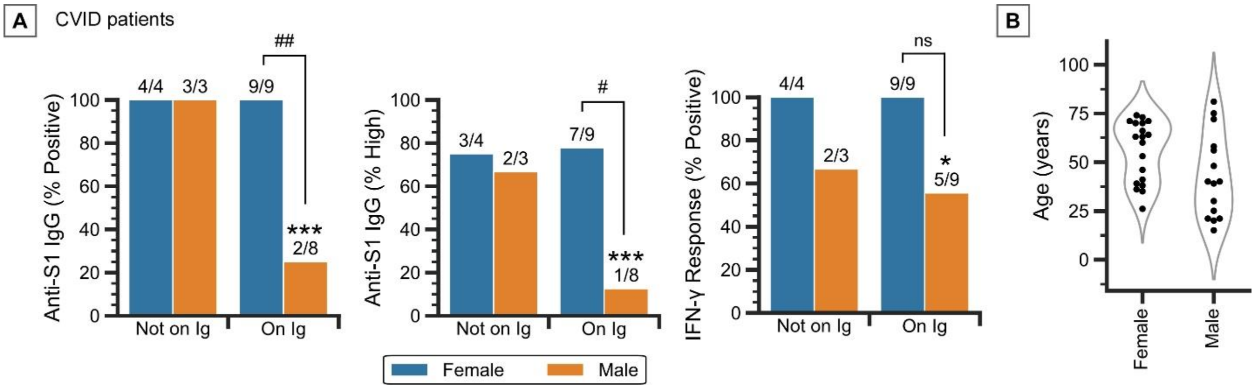

A) Vaccine response positivity rates of CVID patients stratified by immunoglobulin use and gender.

B) Age distribution of male and female patients with immunoglobulin deficiency.

ns, not significant; #*P*<.05, ##*P*<.01, ###*P*<.001 for pairwise comparisons as indicated; **P*<.05, ***P*<.01, ****P*<.001 compared to NISP, Fisher exact test.

### eFigure 8. Vaccine response positivity rates of patients with heme malignancy, stratified by disease subcategory and disease activity.

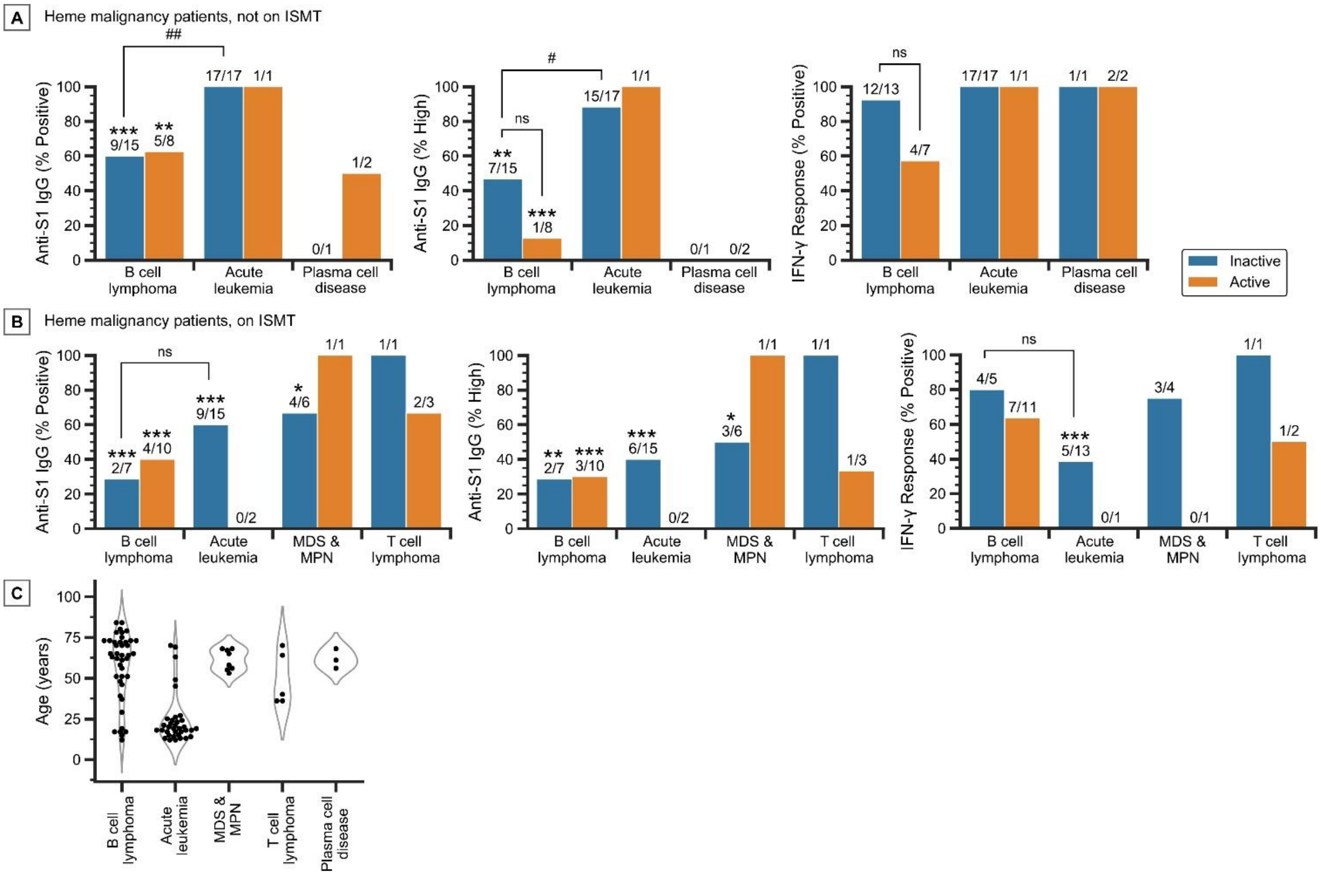

A) Vaccine response positivity rates of patients with heme malignancy and on ISMT.

B) Age distribution of patients with heme malignancy (both on and not on ISMT), stratified by disease subcategory.

Only disease subcategories that apply to three or more patients are plotted. ns, not significant; #*P*<.05, ##*P*<.01, ###*P*<.001 for pairwise comparisons as indicated; **P*<.05, ***P*<.01, ****P*<.001 compared to NISP, Fisher exact test.

**eFigure 9. ISPs show a similar rate of decline in vaccine response over time after primary vaccination as HCWs and NISPs.**

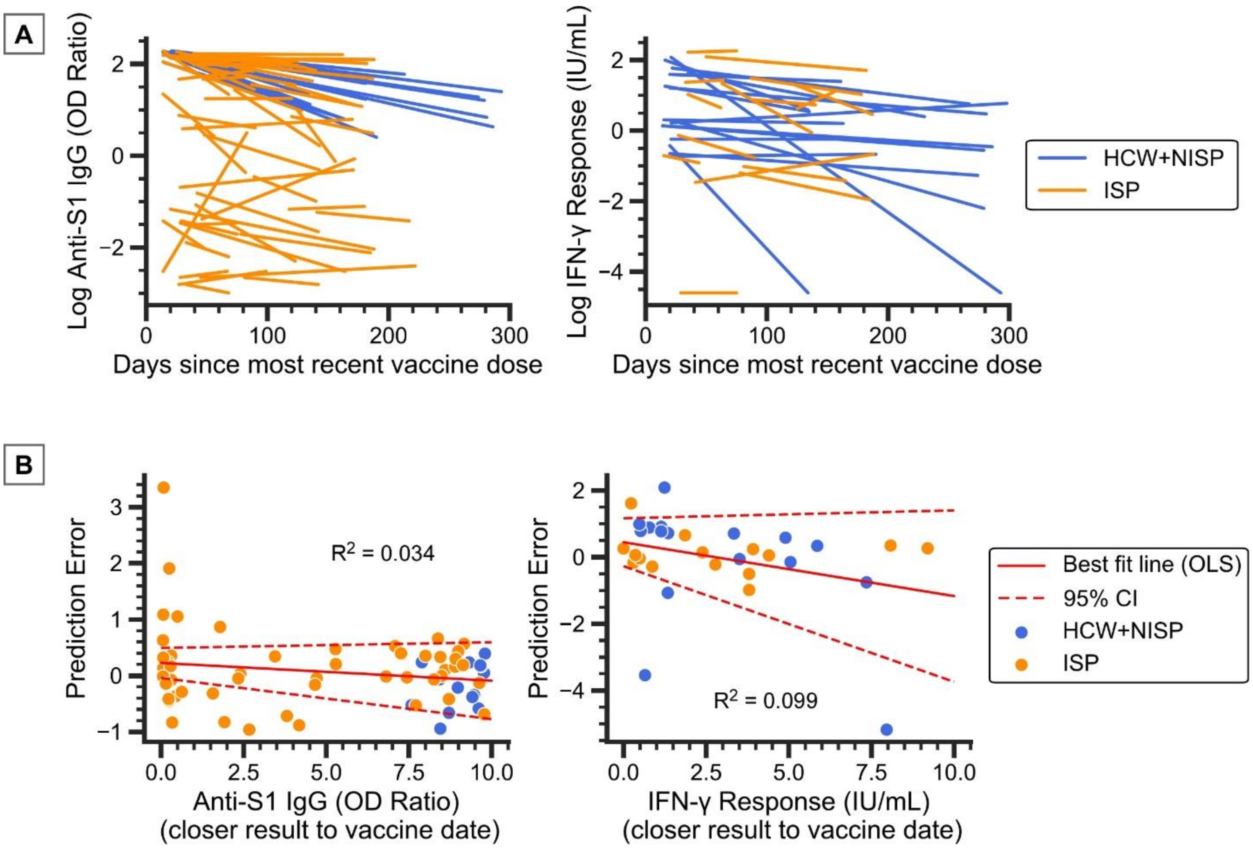

A) Tracking the change in natural log transformed vaccine response over time after primary vaccination in patients with repeated assays.

B) Error (actual minus expected natural log transformed anti-S1 IgG and IFN-γ response) produced by equations 2 and 3 (eMethods) when used to predict vaccine response at different times after primary vaccination. Linear models were fitted to the prediction error versus the initial vaccine response (using the value of the assay performed closest to the vaccination date). Dotted lines denote the 95% confidence interval (CI) of the linear model mean and intercept.

For anti-S1 IgG, n = 12 HCWs, 2 NISPs, 52 ISPs; for IFN-γ response, n=15 HCWs, 1 NISP, 15 ISPs.

### eFigure 10. Determination of anamnestic booster response.

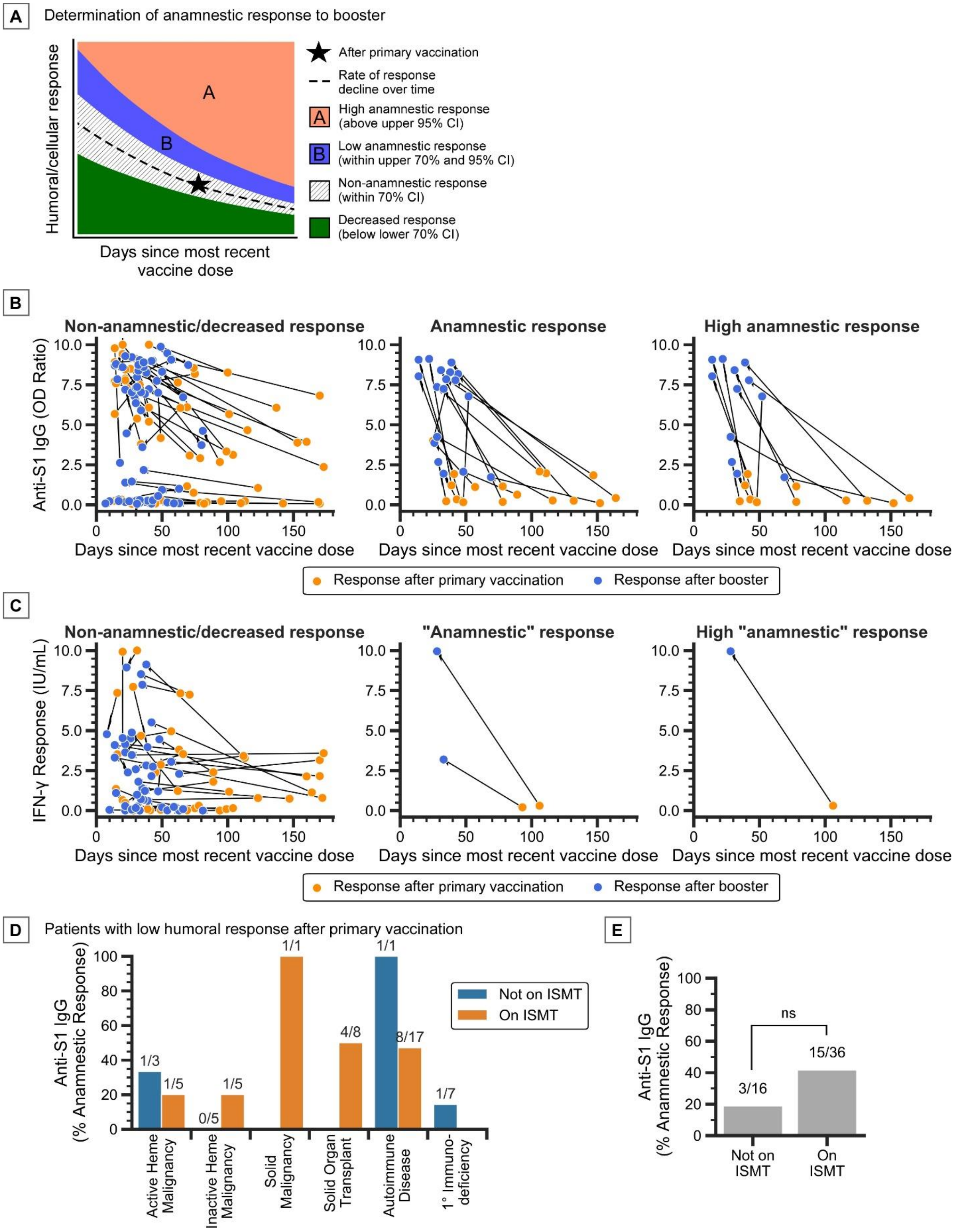

A) Determination of anamnestic booster response given primary vaccination response (Star), based on the expected rate of change in response over time (dashed line), with confidence intervals (CI). Booster responses in sections labeled A and B are determined to be anamnestic. Booster responses in section B (above the upper bound of the 70% CI) is determined to be “low” anamnestic, while booster responses in section A (above the upper bound of the 95% CI) is determined to be “high” anamnestic.

B) HCW (n = 4), NISP (n = 1), and ISP (n = 79) cohorts with IgG assay results both before and after receiving the booster vaccine, separated into non-anamnestic/decreased booster response (n = 4 HCWs, 1 NISPs, 61 ISPs), anamnestic booster response (n = 18 ISPs), and high anamnestic response (n = 12 ISPs).

C) HCW (n = 6) and ISP (n = 42) cohorts with IGRA results both before and after receiving the booster vaccine, separated into non-anamnestic/decreased booster response (n = 6 HCWs, 40 ISPs), anamnestic booster response (n = 2 ISPs), and high anamnestic response (n = 1 ISP).

D) Anamnestic booster response rates in patients stratified by disease category and therapy status. There were six patients with diseases belonging to multiple categories, which we categorized into one single category based on their primary disease for which they were undergoing active therapy/surveillance.

E) Anamnestic booster response rates in patients stratified by therapy status; ns, not significant, Fisher exact test.

D,E: only patients with low anti-S1 IgG after primary vaccination are included.

**eFigure 11. Graphical representation of non-anamnestic vs. anamnestic humoral booster response in patients affected by select immunosuppressive factors.**

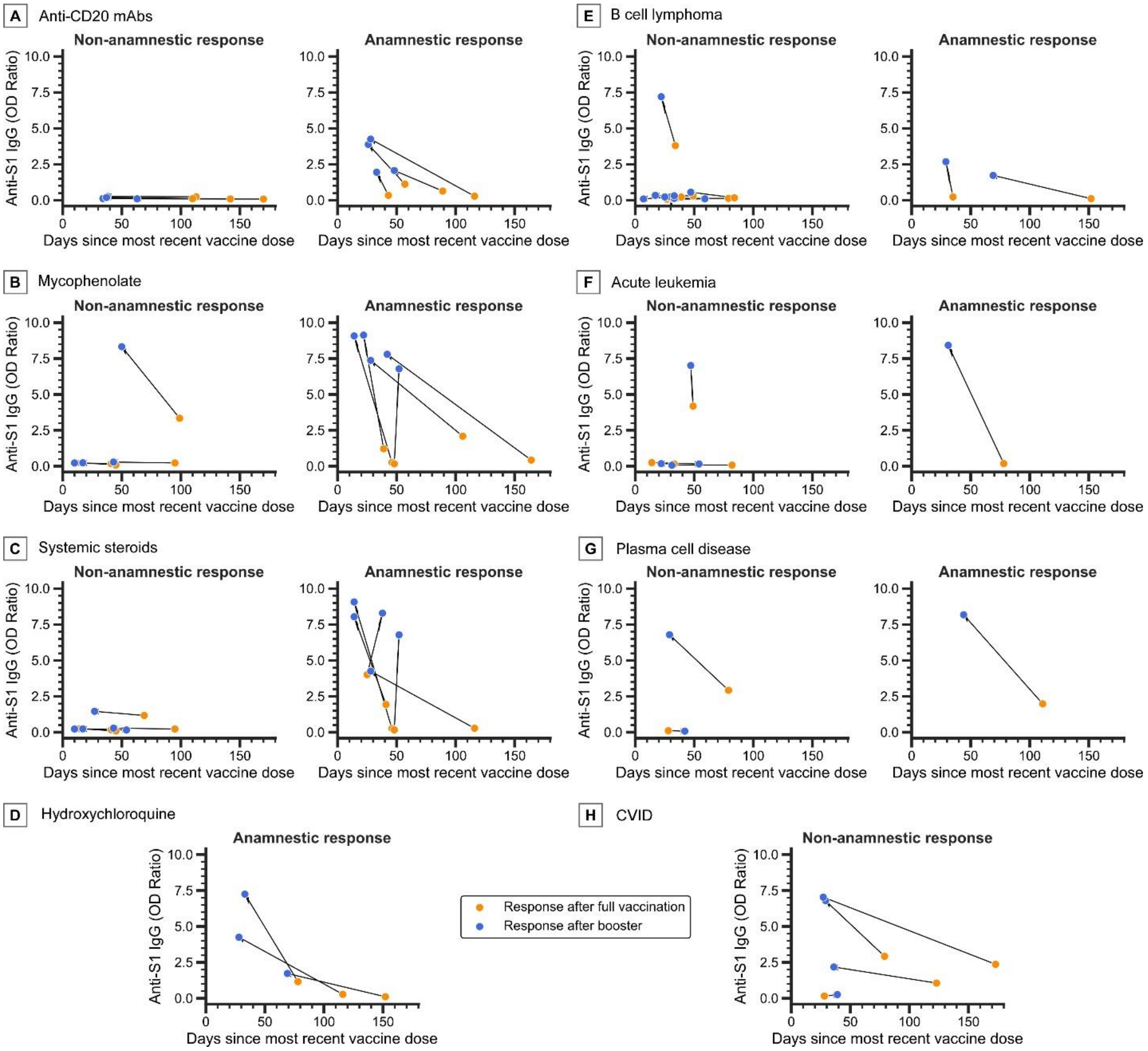

A) Anti-CD20 mAbs (n=4 non-anamnestic, n=4 anamnestic)
B) Mycophenolate (n=4 non-anamnestic, n=5 anamnestic)
C) Systemic steroids (n=5 non-anamnestic, n=5 anamnestic)
D) Hydroxychloroquine (n=0 non-anamnestic, n=3 anamnestic)
E) B cell lymphoma (n=9 non-anamnestic, n=2 anamnestic)
F) Acute leukemia (n=4 non-anamnestic, n=1 anamnestic)
G) Plasma cell disease (n=2 non-anamnestic, n=1 anamnestic)
H) CVID (n=4 non-anamnestic, n=0 anamnestic)

Only shown are ISPs with low anti-S1 IgG after primary vaccination.
